# In vivo mapping of striatal neurodegeneration in Huntington’s disease with Soma and Neurite Density Imaging

**DOI:** 10.1101/2025.03.17.25324107

**Authors:** Vasileios Ioakeimidis, Marco Palombo, Chiara Casella, Lucy Layland, Carolyn B. McNabb, Robin Schubert, Philip Pallmann, Monica E. Busse, Cheney J. G. Drew, Sundus Alusi, Timothy Harrower, Jane Davies, Anne E. Rosser, Claudia Metzler-Baddeley

**Affiliations:** Cardiff University Brain Research Imaging Centre (CUBRIC), School of Psychology, Cardiff University, _Maindy Road_, _Cathays_, _Cardiff_, UK; Danish Research Centre for Magnetic Resonance, Department for Radiology and Nuclear Medicine, Copenhagen University Hospital Amager and Hvidovre, Copenhagen, Denmark; Department for Forensic and Neurodevelopmental Sciences, Institute of Psychiatry, Psychology and Neuroscience, King’s College London, London, UK; Early life imaging research department, School of Biomedical Engineering and Imaging Sciences, King’s College London, London, UK; London Collaborative Ultra high field System (LoCUS), Kings College London, London, UK; George Huntington Institut (GHI), _Wilhelm-Schickard-Strasse 15_, _48149 Muenster_, Germany; Centre for Trials Research, School of Medicine, Cardiff University, _Heath Park Way_, _Cardiff_, UK; The Walton Centre for Neurology and Neurosurgery, _Lower Lane_, _Fazakerley_, Liverpool, UK; Royal Devon and Exeter NHS Trust, _Barrack Road_, _Exeter_, UK; Cardiff and Vale University Health Board, Main University Hospital Wales Building, Cardiff University, _Health Park Campus_, _Cardiff_, UK; Cardiff University Brain Repair Group, School of Biosciences, Cardiff University, _Museum Avenue_, _Cardiff_, UK; Advanced Neurotherapeutics Centre (ANTC), Department of Neurology and Psychological Medicine, School of Medicine, Cardiff University, _Hadyn Ellis Building_, _Cardiff_, UK

**Keywords:** Huntington’s disease (HD), basal ganglia, grey matter microstructure, Soma and Neurite Density Imaging (SANDI), ultra-strong gradient diffusion-weighted MRI, motor performance

## Abstract

**Background:** Huntington’s Disease (HD) is an inherited neurodegenerative disorder characterised by progressive cognitive and motor decline resulting from atrophy within basal ganglia networks. Although no disease-modifying therapies currently exist, several novel clinical trials are ongoing. Sensitive non-invasive imaging biomarkers are therefore essential for evaluating therapeutic effects. Soma and Neurite Density Imaging (SANDI), a multi-shell diffusion-weighted imaging model, estimates intracellular signal fractions arising from sphere-shaped soma that show promise as proxies for HD-related neurodegeneration. Although HD is rare, it offers a valuable model for understanding other neurodegenerative diseases due to its clear genetic cause and shared patterns of protein abnormalities.

**Objective:** To characterise HD-related microstructural abnormalities in the basal ganglia and thalami using SANDI and examine associations between SANDI indices, volumetric measurements, and motor performance.

**Methods:** T1-weighted anatomical and multi-shell diffusion-weighted images (b-values: 200 s/mm²– 6,000 s/mm²) were acquired using a 3T Siemens Connectom scanner (300mT/m) in 56 HD individuals (Mean_Age_ = 46.1, SD_Age_ = 13.8, 25 females) and 57 healthy controls (Mean_Age_ = 45.0, SD_Age_ = 13.8, 31 females). HD participants completed Quantitative Motor (Q-Motor) tasks, including speeded and paced finger tapping, which were reduced to one principal component of motor performance. Following standard diffusion-weighted data preprocessing, SANDI and diffusion tensor models estimated apparent soma density, apparent soma size, apparent neurite density, extracellular signal fraction, fractional anisotropy, and mean diffusivity. The caudate, putamen, pallidum, and thalamus were segmented bilaterally, and micro-structural and volumetric indices were extracted and compared. Correlations between SANDI indices, Q-Motor performance, and volumetric measures were analysed.

**Results:** HD was associated with reduced apparent soma density (*r_rb_* = 0.32, *p* ≤ 0.007) and increased apparent soma size (*r_rb_* = 0.45, *p* < 0.001) and extracellular signal fraction (*r_rb_* = 0.34, *p* ≤ 0.003) in the basal ganglia, but not the thalami. These differences were more pronounced at HD-Integrated Staging System 0-1 than 2-3. No differences were found in apparent neurite density (*r_rb_* = 0.18, *p* = 0.17). HD-related increases in fractional anisotropy and mean diffusivity in the basal ganglia were replicated. Q-Motor component scores correlated negatively with apparent soma density and positively with apparent soma size and extracellular signal fraction. SANDI indices and age explained up to 63% of striatal atrophy in HD.

**Conclusion:** SANDI measures detected HD-related neurodegeneration in the striatum, accounted significantly for striatal atrophy, and correlated with motor impairments. Reduced apparent soma density and increased apparent soma size align with *ex vivo* evidence of medium spiny neuron loss and glial reactivity. SANDI shows promise as an *in vivo* biomarker and surrogate outcome measure for clinical trials of disease-modifying therapies for HD and other neurodegenerative diseases.

## Introduction

Huntington’s disease (HD) is an autosomal dominantly inherited neurodegenerative disorder caused by a pathogenic CAG repeat expansion of the Huntingtin gene.^1^ HD is characterised by a progressive loss of cognitive and motor functions as well as psychiatric disturbances.

The clinical onset of HD is commonly defined by the manifestation of motor symptoms, such as chorea, reduced voluntary motor control, bradykinesia, and difficulty maintaining rhythmic and paced movements.^2–4^ However, changes in the brain, notably striatal atrophy in the basal ganglia (BG)^5,6^ may precede the motor onset by up to 24 years^7–10^ and correlate with motor and cognitive decline.^2,5,6,11–15^ While HD is rare (∼12 in 100,000), it can be seen as a model neurodegenerative disorder, due to its clear genetic cause, well-characterized disease progression, and shared features of protein abnormalities with more common disorders like Alzheimer’s and Parkinson’s disease.^5^

There is presently no approved disease-modifying therapy for HD, but several clinical trials are underway to test the safety and efficacy of novel therapeutics.^16^ The recent surge in potential disease-modifying targets has generated a demand for surrogate outcome measures that are sensitive to HD neuropathology and allow a mechanistic assessment of therapeutic effects on striatal neurodegeneration in a timely manner. Volumetric measurements from non-invasive MRI are known to be sensitive to disease progression^17–20^ and have been adopted into the Huntington’s Disease Integrated Staging System (HD-ISS).^21^ However, volumetric measurements do not provide information about the underlying neuropathological tissue changes that lead to striatal atrophy, such as the loss of medium spiny neurons (MSN)^22,23^ and changes in glia cell density and morphology, including enlargement of reactive astrocytes and microglia.^22,24–26^

Diffusion-weighted Imaging (DWI) is widely used to investigate brain tissue microstructure *in vivo* by exploiting apparent water displacement due to Brownian motion.^27,28^ Most DWI studies in HD have used diffusion tensor imaging (DTI), which models extra-cellular water diffusion as a Gaussian tensor and measures diffusion properties such as mean diffusivity (MD) and the degree of diffusion anisotropy (fractional anisotropy; FA).^29^ DTI studies in HD have consistently reported increases in MD and FA in striatal grey matter,^30,31^ that likely arise from the selective neurodegeneration of MSN connections.

Advances in multi-shell and ultra-strong gradient DWI,^32^ have enabled increasingly sophisticated biophysical models that require data acquisition over a range of b-values to separate extra- from intracellular diffusion signals.^33–35^ Several approaches that model intraneurite space with cylinders or sticks have been put forward (e.g. Composite and Restricted Model of Diffusion, CHARMED^33^; Convex optimization modelling for microstructure informed tractography; COMMIT^36^; Neurite Orientation Dispersion and Density Index; NODDI^34^). NODDI studies in premanifest HD have revealed reduced apparent neurite density in white mater with localised reductions in fibre orientation dispersion in the corpus callosum and basal ganglia capsules.^37^ However, despite putamen volume loss, no NODDI-based differences in striatal grey matter were detected in gene-positive individuals long before motor onset.^10^

Soma And Neurite Density Imaging (SANDI)^38^ is a diffusion MRI model that extends biophysical multi-compartment models to capture microstructural features of cell bodies and processes *in vivo*. SANDI requires multi-shell acquisition protocols with b-values over 3,000 s/mm^2^ to capture intracellular restricted signal fractions. Biologically, its parameters describe the relative contributions of somas, neurites (axons, dendrites and glial processes), and extracellular space to the diffusion-weighted MRI signal: the soma signal fraction (f_is_) reflects the density of neuronal and glial cell bodies, the neurite fraction (f_in_) captures density of axons, dendrites and glial processes, and the extracellular fraction (f_extra_) and extracellular diffusivity (D_e_) account for the surrounding milieu. The MR apparent soma radius (r_s_) further provides an estimate of (volume-weighted) average cell body size. Mathematically, SANDI models the diffusion signal attenuation as a weighted combination of restricted diffusion in spheres (somas), restricted diffusion in cylinders of infinitesimally small radius (neurites), and Gaussian diffusion in the extra-cellular compartment, constrained by the condition that the compartment fractions sum to one. These parameters thus provide biologically interpretable and mathematically well-defined indices of cellular composition, enabling non-invasive characterization of brain microstructure beyond what can be achieved with conventional diffusion models.

SANDI has been shown to provide highly reproducible and repeatable parameter estimation across grey matter regions in the human brain^39^ that align closely with its known cyto- and myeloarchitecture. For instance, the gradients of apparent soma density maps were found to closely match those of Brodmann areas in human cortical regions with different soma density profiles,^38^ and correlated in the mouse brain with cell density distributions from the Allen atlas.^38,40^ These findings suggest the potential of the SANDI model for quantifying neurodegenerative processes in the grey matter of the living human brain.

Clinical applications of SANDI have so far been limited to multiple sclerosis (MS) and amyotrophic lateral sclerosis (ALS). In MS, reductions in apparent soma and neurite density, together with increases in extracellular signal fraction, have been reported in both grey and white matter, consistent with demyelination, axonal loss, and neurodegeneration.^41,42^ These abnormalities correlated with disease severity,^42,43^ cortical and subcortical atrophy,^41,44^ and elevated serum neurofilament light chain levels, a marker of axonal damage^44^ that is also sensitive to HD progression.^45,46^ Similar patterns, namely reduced apparent soma density and region-specific alterations in apparent soma size have recently been observed in ALS, ^43^ aligning with motor neuron loss and glial responses.

The primary objective of this study was to determine whether SANDI indices were sensitive and specific to microstructural grey matter differences in the BG, compared with the thalami, in individuals with HD relative to healthy controls. The thalami were selected as control regions based on the established trajectory of neurodegeneration in HD, which begins with early loss of MSN in the striatum before extending to neighbouring structures such as the putamen and thalamus. At most participants were at early disease stages, we assumed the thalami would remain relatively unaffected in this sample.

Secondary objectives were to explore the extent to which HD-related SANDI differences accounted for BG atrophy, assessed using volumetric measurements, and for performance differences in motor tasks, including speeded and paced finger tapping, which are known to be associated with striatal atrophy in HD. ^47^ Finally, relationships between SANDI indices and disease burden using the HD-ISS ^21^ and the CAG-Age Product (CAP_100_) score ^48^ were explored.

## Materials and methods

### Participants

MRI data from 56 individuals with HD and 57 age- and sex-matched healthy controls (HC) were included in the analyses. Imaging data were retrospectively pooled from three projects, all of which acquired scans on the same Siemens Connectom system at the Cardiff University Brain Research Imaging Centre (CUBRIC) using identical acquisition protocols. Thirty-eight of the HD participants were drawn from a randomised controlled feasibility trial of HD-DRUM,^49^ a remote rhythmic training intervention, with ethical approval from the Wales Research Ethics Committee 2 (REC Reference: 22/WA/0147).^55^ Here we report their baseline MRI and behavioural data collected prior to randomisation. An additional 18 individuals with HD and 18 age-matched HC were included from a previous study of white matter microstructure in premanifest HD (REC Reference: 18/WA/0172).^50^ Further comparison data were obtained from 25 age- and sex-matched HC from the Wales Advanced Neuroimaging Database (WAND) Study^51^ (REC Reference: 18.08.14.5332RA3) and 14 HC from the HD-DRUM trial. All participants provided written informed consent in accordance with the Declaration of Helsinki.

HD individuals were identified and screened for eligibility in five clinics in the UK (Bristol, Birmingham, Cardiff, Exeter, and Liverpool). HC volunteers were recruited from online advertisements on the Cardiff University social network, Viva Engage, or in HD clinics as support partners or family members. HC participants were also recruited through [Healthwise Wales](https://www.healthwisewales.org/) and by word of mouth. For the WAND study, data collection was reported elsewhere.^51^

Individuals aged 18 years or older with a good command of English were eligible to participate. Additional inclusion criteria for individuals with HD^49^ were a positive genetic test for the presence of the mutant huntingtin allele (CAG length ≥ 36 repeats) and/or clinical diagnosis of HD, along with a Unified Huntington’s Disease Rating Scale (UHDRS) Total Functional Capacity (TFC) score between 9 and 13.^52^ Exclusion criteria for all participants included an inability to provide informed consent and any contraindication for MRI (e.g. pacemakers, stents). Further exclusion criteria were a history of any other neurological condition for HD participants, and for HC, a history of neurological or psychiatric disorders, and/or alcohol or drug abuse associated with grey matter volume loss.

To characterise general cognitive functioning, HD participants completed the Montreal Cognitive Assessment (MOCA).^53^ Verbal intellectual ability was assessed using the Test of Premorbid Functioning (TOPF).^54^ Disease burden was estimated by the TFC score and the CAG-Age Product (CAP100) score,^48^ calculated as:

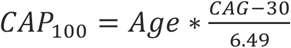

HD-ISS^21^ staging and associated clinical information for stratification were obtained from the Enroll-HD^55^ observational study (former Registry; REC no 04/WSE05/89). However, due to the retrospective pooling of datasets, HD-ISS information was available for only 30 HD participants (see Supplementary Table 1 for demographic and clinical characteristics).

### Motor outcome measures

Participants in the HD-DRUM study completed a range of motor tasks from the Quantitative-Motor (Q-Motor) test-battery,^56–58^ which provided reliable assessments of speeded finger tapping performance in clinical HD trials.^59,60^ Tasks included left and right 1) speeded index finger and foot tapping using force transducers,^47^ 2) paced finger and foot tapping^47^ with a metronome-paced and memory-paced phase, using a fast (0.55s inter-onset interval; IOI) or slow (1.1s IOI) pace, 3) 3D pointing to four target locations in a predefined sequence using a position-tracking stylus with the dominant hand^61^ and 4) 3D target pointing and speeded finger tapping dual task performed with dominant and non-dominant hand, respectively. Outcome measures included mean IOI (seconds) and area under the curve (AUC) (Newton-seconds) for the speeded tapping, mean absolute deviation from the metronome pace (seconds) for the paced tapping, target frequency (Hz) for the target pointing, and target frequency, mean IOI and AUC for the dual-task condition.

### Image acquisition

All MRI data included in the analyses were acquired on the same 3T Siemens Connectom scanner (Siemens Healthcare, Erlangen, Germany) with ultra-strong magnetic gradients (300mT/m) at CUBRIC using identical acquisition protocols as detailed below. No scanner changes or upgrades were performed during the data collection periods.

T1-weighted (T1w) images were acquired using a magnetisation-prepared 180-degrees radio-frequency pulses and rapid gradient-echo (MPRAGE), with the following parameters: repetition time (TR) 2,300 ms, echo time (TE) 2 ms, field of view (FOV) 256 x 256 x 192 mm, matrix size 256 x 256 x 192, resolution 1 x 1 x 1 mm^3^, flip angle 9°, inversion time (TI) 857 ms, in-plane acceleration (GeneRalised Autocalibrating Partial Parallel Acquisition; GRAPPA) factor 2, phase-encoding direction anterior to posterior (AP), and acquisition time of 6 minutes.

Multi-shell High Angular Resolution Diffusion Imaging (HARDI)^62^ data were obtained at b-values of 200 s/mm² (20 directions), 500 s/mm² (20 directions), 1,200 s/mm² (30 directions). 2,400 s/mm² (61 directions), 4,000 s/mm² (61 directions) and 6,000 s/mm² (61 directions) using a single-shot spin-echo, echo-planar imaging sequence with TR = 3,000 ms, TE = 59 ms, FOV 220 x 200 mm in-plane; matrix size 110 x 110 x 66; 2 mm^3^ resolution, gradient pulse duration - δ = 7 ms, gradient pulses separation - Δ = 24 ms in AP phase-encoding direction with an in-plane acceleration (GRAPPA) factor of 2. Fifteen non-diffusion-weighted (b-value = 0 s/mm²) images were acquired [two initial and 11 interspersed at the 33^rd^ volume and every 20^th^ volume thereafter in AP direction and 2 images in the posterior-to-anterior (PA) direction]. The HARDI acquisition time was 18 minutes.

### Image processing

#### Diffusion-weighted image preprocessing

Multi-shell HARDI data were pre-processed and corrected for signal drift, susceptibility-induced distortions, motion and eddy current-induced distortions, gradient non-uniformity and Gibbs ringing artifacts using a custom in-house pipeline comprising tools from the FMRIB Software Library (FSL version 6.0.3),^63^ the MRtrix software package,^64^ ExploreDTI^65^ (version 4.8.6) and in-house MATLAB-based scripts.^39^

The FSL brain extraction tool^63^ was used to mask the first non-diffusion-weighted image from each phase-encoding direction to exclude non-brain data. The diffusion-weighted MRI volumes were fitted to temporally interspersed b0 volumes to correct for within-image intensity drift by using custom code in MATLAB R2017b (MathWorks Inc., Natick, Massachusetts, USA). Slicewise outlier detection (SOLID)^66^ was applied with modified Z-score thresholds of 3.5 (lower) and 10 (upper), utilising a variance-based intensity metric. FSL’s top-up tool^67,68^ was used to estimate susceptibility-induced off-resonance fields from b0 images that were acquired in opposing phase-encoding directions (AP and PA) and then FSL’s eddy tool^69^ was used to correct eddy current-induced distortions and subject movements. Gradient non-uniformity distortions were corrected using in-house code in MATLAB R2017b. Finally, Gibbs ringing correction was performed in MRtrix3 using the local subvoxel-shifts method.^70^

For the purpose of comparing our results with the previous literature,^30^ DTI was fitted with ExploreDTI using data with b-values of 500 s/mm² and 1,200 s/mm² to produce outcome maps for FA and MD, estimated with linearly weighted least squares regression.

#### SANDI analysis

The SANDI model ^38^ assumes three compartments, namely intra-neurite signal modelled as diffusion inside impermeable randomly oriented sticks, intra-soma signal modelled as restricted diffusion inside spheres, and extra-cellular signal modelled as Gaussian isotropic diffusion. The direction-averaged (or spherical mean) normalized diffusion signal has thus the following expression:

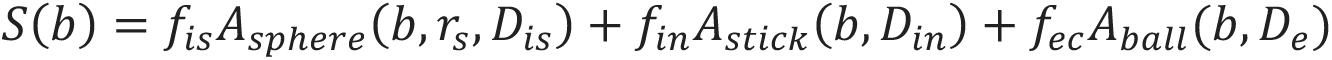

where f_in_ + f_is_ + f_ec_ = 1; A_stick_ and A_sphere_ are the normalized, directionally-averaged (or spherical mean) signals for restricted diffusion within neurites and soma, respectively and A_ball_ is the normalized, directionally-averaged (or spherical mean) signal of the extra-cellular space. The specific expressions are given in ^38^.

The parameters estimated from the direction-averaged (or spherical mean) data are D_in_, proxy of the intra-neurite effective axial diffusivity; D_e_, proxy of the extracellular effective mean diffusivity; r_s_, a proxy of apparent soma radius as well as the signal fractions subject to the constraint f_in_ + f_is_ + f_ec_ = 1, proxy respectively of the relaxation-weighted neurite, soma and extracellular volume fractions. The bulk diffusivity inside the sphere D_is_ is fixed to 3 μm^2^/ms.

The parameters were fitted using a Random Forest regression algorithm (TreeBagger Matlab®) with 200 trees, trained on simulated data, using the code publicly available at https://github.com/palombom/SANDI-Matlab-Toolbox-Latest-Release. The training data consisted of simulated signals for 10^5^ parameter combinations, uniformly sampled: f_in_ and f_is_ ∈ [0, 1], D_in_ ∈ [0.5, 3] μm^2^/ms, D_e_ ∈ [0.5, 3] μm^2^/ms and r_s_ ∈ [1, 12.5] μm. Rician noise with a distribution of standard deviations randomly sampled from the voxels within the brain mask of the noise map was obtained using Marchenko-Pastur principal component analysis (MP-PCA)-based method^71–73^ in MRtrix3 and was added to account for realistic SNR levels and rectified noise floor. The loss function of the training was the mean squared error between predicted parameters and ground truth values. These noise maps were subsequently used to fit the SANDI model^38^ to the pre-processed multi-shell diffusion data with the SANDI MATLAB Toolbox (https://github.com/palombom/SANDI-Matlab-Toolbox-Latest-Release^38^ using all the default settings.

The model fitting produced maps of the intra-neurite, extracellular and intra-soma signal fractions (f_in_, f_ec_, f_is_), apparent soma size (r_s_; measured in μm) and intra- and extra-neurite diffusivities (D_in_, D_e_; measured in mm^2^/ms). Post-hoc sensitivity analysis of the SANDI model parameters revealed very low sensitivity to changes in D_in_. Consequently, it was excluded from further analysis.

#### T1-weighted image preprocessing

The default FreeSurfer ^74^ (v6) *recon-all* pipeline was utilised to segment subcortical BG ROIs of the caudate, putamen, and globus pallidum as well as of the thalamus as control ROIs. ROIs were segmented from T1w images and were identified and labelled in each hemisphere.

#### Extraction of microstructural metrics from regions-of-interest

Median values of each microstructural index from DTI (FA, MD) and SANDI (f_is_, f_in_, f_ec_, r_s_, D_e_) models were extracted for each ROI using FSL’s *fslmaths.* ROI masks were aligned with the diffusion space using rigid transformation with FSL’s *flirt*^75^ before eroding the boundaries of the subcortical masks by one voxel with the default 3 × 3 × 3 kernel settings to minimise partial volume effects and then aligning all microstructural maps with the masks.

Volumetric measures for each ROI and intra-cranial volume (ICV) were extracted from FreeSurfer v6 ROI volumes were normalised for ICV. The addition of brain volumes allowed exploration of the extent to which any HD-related SANDI differences accounted for BG atrophy.

### Statistical analysis

Statistical analyses were performed in JASP (v0.18.1.0)^76^, R version 4.4.1 (2024-06-14)^77^ in R-studio (2024.9.0.375)^78^ and SPSS (v27) (IBM Corp).^79^ Data normality was assessed using the Shapiro-Wilk test, with p < 0.05 indicating non-normal distribution. Descriptive statistics for each group were reported as percentages (%), means and standard deviations (SD). Medians of each microstructural index in each ROI were compared between the groups with Mann-Whitney-U tests because of lack of normality and unequal variance between groups. Effect sizes (ES) for group comparisons were therefore reported with rank biserial correlation (*r_rb_*). Multiple comparisons were corrected with Benjamini-Hochberg’s method to control a false discovery rate (FDR) of 0.05^80^ and applied to all statistical tests that related to the same theoretical inference.^81^

Multi-collinearity of imaging metrics was explored by calculating the intercorrelations between all SANDI, DTI, and volumetric measurements averaged across all four ROIs (caudate, putamen, pallidum, and thalamus). Spearman’s rho (*ρ*) correlation matrices were calculated in the full sample, as well as the HC and HD groups separately.

Hierarchical linear regression analyses were conducted to explore SANDI predictors of the variance in volumetric measurements. Regression analyses were carried out for each ROI and each group separately. HD data were modelled by firstly accounting for age and TFC scores (available for all HD participants) simultaneously. This was followed by step-wise inclusion of all SANDI indices using an iterative forward selection and backward elimination method based on each variable’s F-statistic and p-value that aimed to maximise the adjusted R^2^-value while keeping only the most significant predictors. HD and HC data were modelled in the same way using all SANDI variables and age, except for the inclusion of TFC scores that were only available for HD participants.

Principal Component Analysis (PCA) was carried out to reduce the dimensionality of HD participants’ Q-Motor data and hence the number of multiple correlations with microstructural SANDI indices. PCA followed established guidelines to limit the number of extracted components in relatively small sample sizes.^82,83^ First the Kaiser criterion of including all components with an eigenvalue greater than 1 was applied and the Cattell scree plot was inspected to identify the minimal number of components that accounted for most variability in the data. Each extracted component was then assessed for interpretability. PCA was conducted using orthogonal Varimax rotation of the component matrix with Kaiser normalization. Loadings that were greater than 0.5 were considered to be statistically significant.

Spearman’s rho (*ρ*) correlations were then calculated between HD participants’ motor component scores and the CAP100 with the SANDI indices, DTI, and volumetric measures in each ROI.

HD-ISS categories were obtained either directly from Enroll-HD or calculated using the online HD-ISS calculator (https://enroll-hd.org/calc/html_basic.htm) based on caudate and putamen volumes derived from the FreeSurfer v6^74^ cross-sectional pipeline. For participants labelled “<2” in the Enroll-HD database (reflecting missing imaging data), we distinguished between stages 0 and 1 using two complementary approaches. First, adjusted normative modelling was applied, in which caudate and putamen volumes from healthy controls were modelled using linear regression with age, sex, and ICV as covariates, and individual z-scores were generated from model residuals. Second, raw group-based normalisation was performed by z-scoring ICV-normalised volumes relative to the control mean and SD. Participants labelled “<2” were classified as HD-ISS 1 if either adjusted caudate or putamen z-scores fell ≥ 2 SD below the control mean; otherwise, they were assigned to stage 0. Both approaches produced identical classifications.

Explorative analyses of HD-ISS-related differences in SANDI indices were conducted for BG ROIs. Due to small sample sizes, participants at Stages 0 and 1 were combined (HD-ISS 0-1), as were those at Stage 2 and 3 (HD-ISS 2-3). SANDI metrics for each ROI were averaged across hemispheres. Pairwise comparisons between HD-ISS 0-1 versus HC, HD-ISS 0-1 versus HD-ISS 2-3, and HD-ISS 2-3 versus HC were performed using Mann–Whitney U tests without additional FDR correction.

## Results

### Demographics

Sample characteristics, including age and sex distribution for the HD and HC groups are described in Table 1. The groups were comparable with regards to age and sex (*p*>0.05).

**Table 1.**
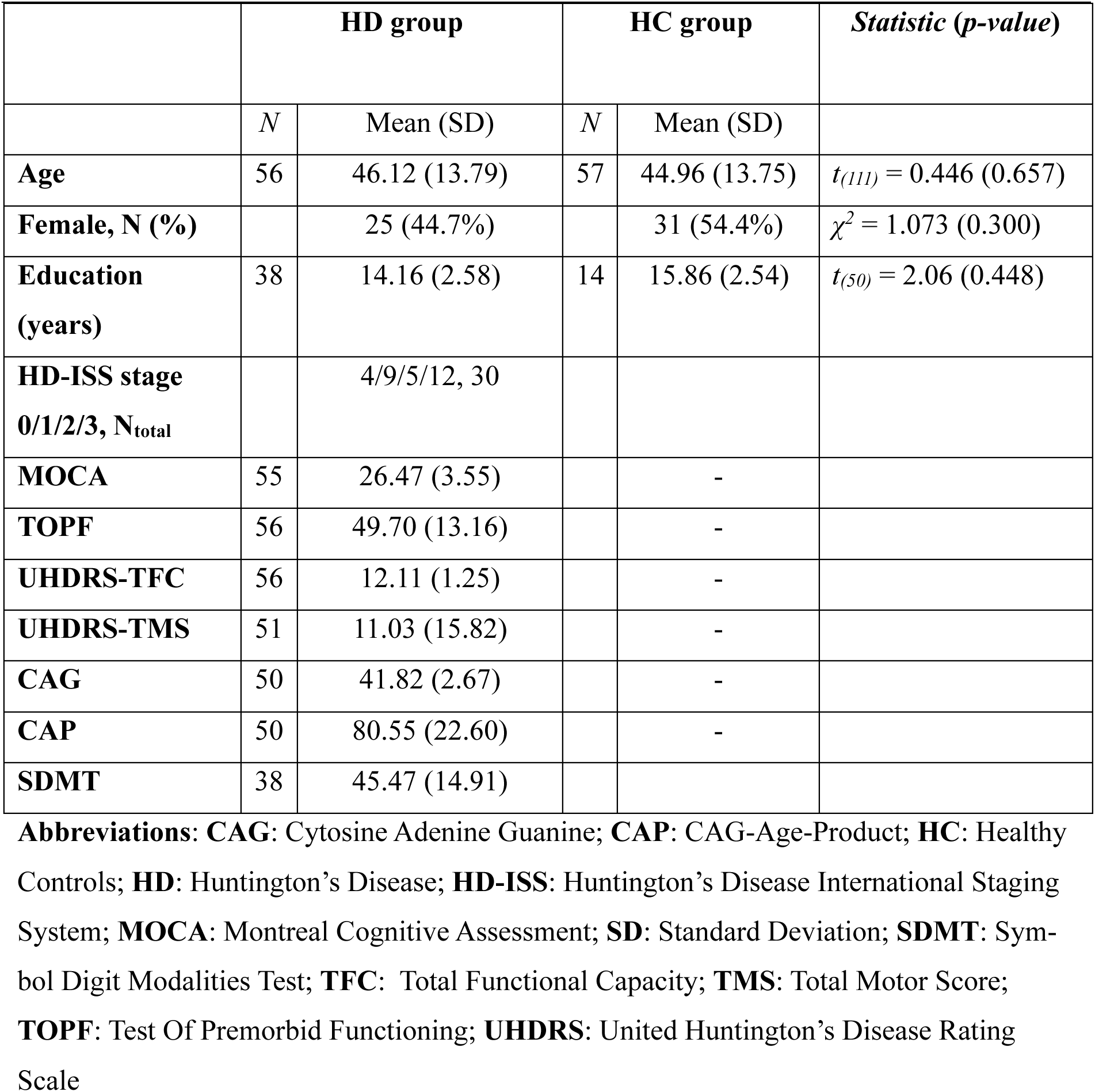
Demographic and clinical information of participants.

Information for HD-ISS calculation was available for 30 individuals. Of these 4 were stratified into Stage 0, 9 into Stage 1, 5 into Stage 2, and 12 into Stage 3; no participants were classified as Stage 4. One individual could not be assigned a stage because their clinical presentation did not align with the HD-ISS assumption that motor symptoms precede functional decline. An additional, 25 individuals could not be classified due to missing clinical data or CAG repeat lengths between 36-39. Demographic and clinical characteristics for each HD-ISS stage, as well as for unclassified participants are summarised in Supplementary Table 1. Supplementary Table 2 provides descriptive statistics for the Q-Motor measures.

### Imaging analysis

#### Differences between HD and HC groups in volumetric measures

Table 2 summarises the volumetric group differences across the eight ROIs with their ES. Figure 1 illustrates the ES maps of volumetric differences between HD and HC, as well as raincloud plots for each ROI. Mann-Whitney tests identified significantly reduced volumes in all BG ROIs in HD compared to HC with moderate ES (*r_rb_* = 0.46-0.55). In contrast, no reductions were present in the right (*FDR-p* = .05) and left thalamus (*FDR-p* = .063) (both *r_rb_* = 0.2).

**Figure 1.**
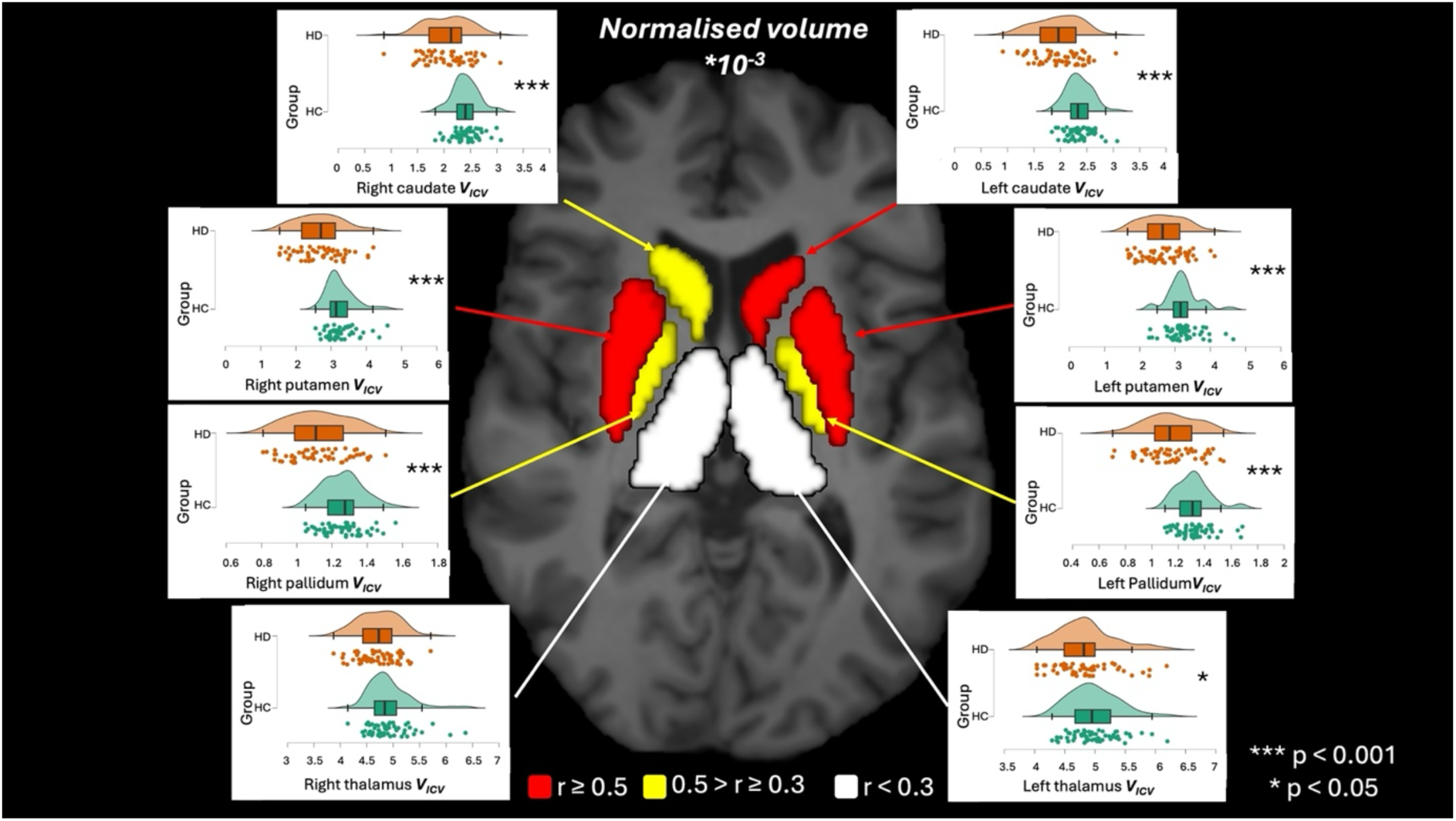
Volumetric differences in the basal ganglia and thalamus between HD and healthy control (HC) groups. Regions-of-interest (ROIs) were segmented using FreeSurfer v6. All ROIs, except the left thalamus, showed significantly smaller volumes in the HD cohort after FDR correction for multiple comparisons. Colours indicate the strength of rank-biserial correlations (*r_rb_*) from Mann-Whitney U tests: Red = strong effect (*r_rb_* ≥ 0.5), Yellow = medium effect (0.3 ≤ *r_rb_* < 0.5), White = small effect (*r_rb_* < 0.3). Raincloud plots show the distribution of the volumetric measures in each ROI per group with orange for HD and green for HC participants. * *p* < 0.05; *** *p* < 0.001

**Table 2.** Descriptive and Mann-Whitney Statistics for Intracranial Volume-Normalised Regions of Interest.

|  |  | **HD group** | **HC group** | **Statistic (FDR-p, effect size)** |
| --- | --- | --- | --- | --- |
| **Region of interest** | **L/R** | **Mean (SD)** | **Mean (SD)** | **U (p, rank-biserial correlation)** |
| *Caudate* | *L* | 1.95<sup>a</sup> (0.45<sup>a</sup>) | 2.35<sup>a</sup> (0.24<sup>a</sup>) | **2464 (<0.001, 0.544)** |
|  | *R* | 2.06<sup>a</sup> (0.43<sup>a</sup>) | 2.41<sup>a</sup> (0.25<sup>a</sup>) | **2376 (<0.001, 0.489)** |
| *Putamen* | *L* | 2.67<sup>a</sup> (0.62<sup>a</sup>) | 3.22<sup>a</sup> (0.46<sup>a</sup>) | **2397 (<0.001, 0.502)** |
|  | *R* | 2.67<sup>a</sup> (0.66<sup>a</sup>) | 3.25<sup>a</sup> (0.40<sup>a</sup>) | **2479 (<0.001, 0.553)** |
| *Pallidum* | *L* | 1.15<sup>a</sup> (0.20<sup>a</sup>) | 1.32<sup>a</sup> (0.13<sup>a</sup>) | **2382 (<0.001, 0.492)** |
|  | *R* | 1.13<sup>a</sup> (0.17<sup>a</sup>) | 1.26<sup>a</sup> (0.12<sup>b</sup>) | **2323 (<0.001, 0.456)** |
| *Thalamus* | *L* | 4.83<sup>a</sup> (0.49<sup>a</sup>) | 4.99<sup>a</sup> (0.41<sup>b</sup>) | 1955 (0.063, 0.224) |
|  | *R* | 4.72<sup>a</sup> (0.39<sup>a</sup>) | 4.91<sup>a</sup> (0.40<sup>b</sup>) | **1981 (0.048, 0.241)** |

**Abbreviations:** **FDR:** False Discovery Rate; **HC:** Healthy Controls; **HD:** Huntington's Disease; **L:** Left Hemisphere; **R:** Right Hemisphere; **SD:** Standard Deviation
<sup>a</sup>: Multiplied by 10<sup>-3</sup>

#### Differences between HD and HC groups in microstructural measures

Descriptive and statistical microstructural results are shown in Table 3. Figure 2 provides ES mappings and raincloud plots of the different microstructural measures for each ROI comparing the two groups.

**Figure 2.**
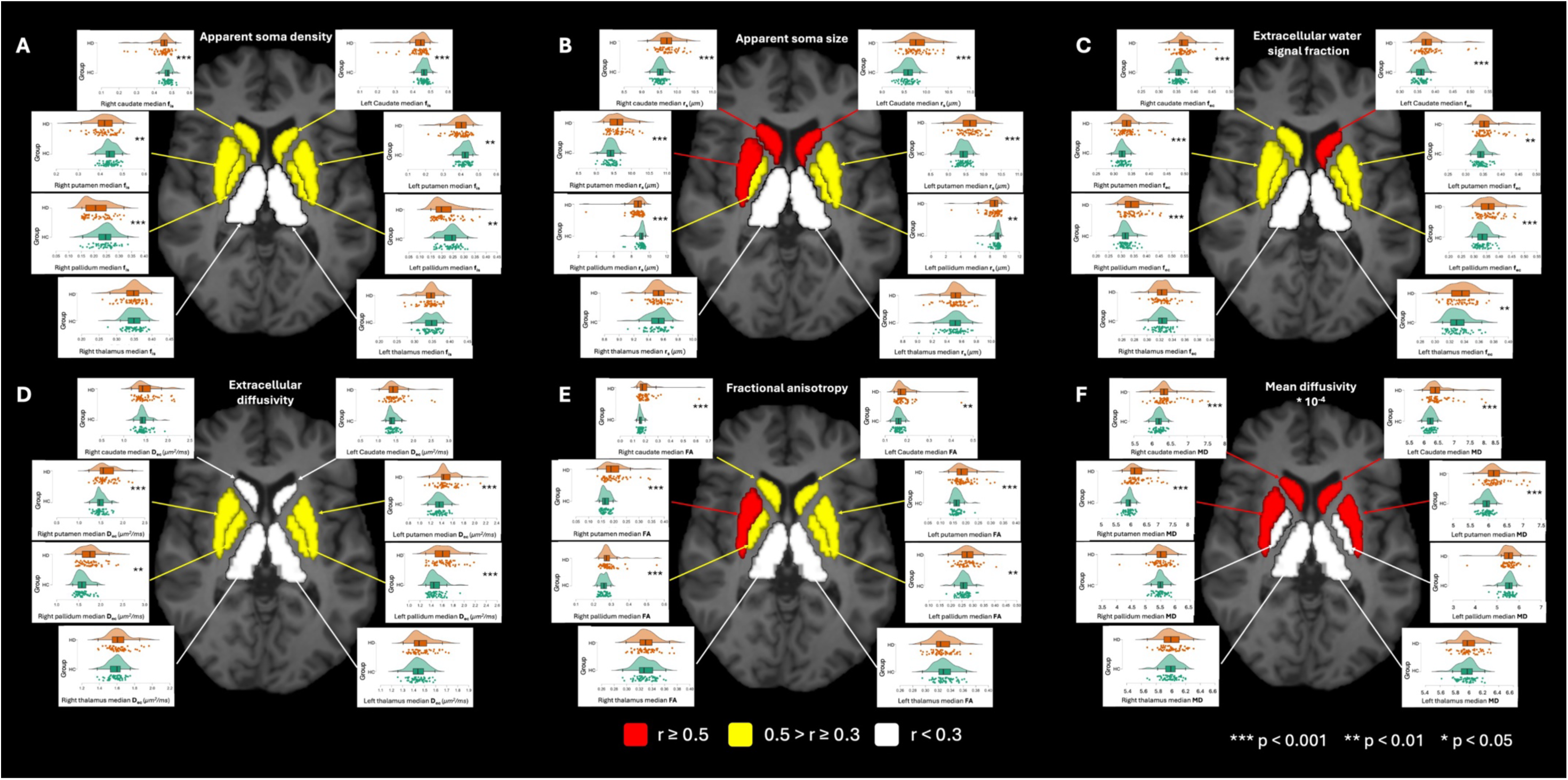
Microstructural differences in ROIs between HD and healthy control (HC) groups. Median values of each microstructural measure were extracted per ROI. **A)** HD individuals show reduced soma density (f_is_) in the basal ganglia (BG). **B)** Soma radius (r_s_) is elevated in the caudate and putamen but reduced in the pallidum. **C)** Extracellular water fraction (f_ec_) is increased in BG regions in the HD group. **D)** Extracellular diffusivity (D_e_) is higher in the putamen and pallidum. **E)** Fractional anisotropy (FA) is elevated in the BG, and **F)** mean diffusivity (MD; expressed in x10^-4^mm²/s) is increased in the striatum. Colours indicate the strength of rank-biserial correlations (*r_rb_*) from Mann-Whitney U tests: Red = strong effect (*r_rb_* ≥ 0.5), Yellow = medium effect (0.3 ≤ *r_rb_* < 0.5), White = small effect (*r_rb_* < 0.3). Raincloud plots show the distribution of the volumetric measures in each ROI per group with orange for HD and green for HC participants. * *p* < 0.05; ** *p* < 0.01; *** *p* < 0.001

**Table 3.** Descriptive and Mann-Whitney Statistics for Microstructural Measures in Regions of Interest.

|  |  |  | **HD group** | **HC group** | **Statistic (FDR-p, effect size)** |
| --- | --- | --- | --- | --- | --- |
| **Microstructural measure** | **Region of interest** | **L/R** | **Mean (SD)** | **Mean (SD)** | ***U* (*p*, *rank-biserial correlation*)** |
| *Apparent soma density* | *Caudate* | *L* | 0.43 (0.05) | 0.46 (0.02) | **2227 (<0.001, 0.395)** |
|  |  | *R* | 0.45 (0.05) | 0.48 (0.02) | **2318 (<0.001, 0.452)** |
|  | *Putamen* | *L* | 0.39 (0.05) | 0.42 (0.03) | **2140 (0.003, 0.341)** |
|  |  | *R* | 0.42 (0.05) | 0.44 (0.03) | **2099 (0.007, 0.315)** |
|  | *Pallidum* | *L* | 0.21 (0.05) | 0.24 (0.04) | **2187 (0.001, 0.370)** |
|  |  | *R* | 0.21 (0.05) | 0.24 (0.04) | **2249 (<0.001, 0.409)** |
|  | *Thalamus* | *L* | 0.34 (0.03) | 0.34 (0.02) | 1692 (0.701, 0.060) |
|  |  | *R* | 0.35 (0.03) | 0.35 (0.02) | 1661 (0.776, 0.041) |
| *Apparent soma size* | *Caudate* | *L* | 9.80 (0.25) | 9.58 (0.15) | **746 (<0.001, -0.533)** |
|  |  | *R* | 9.72 (0.25) | 9.52 (0.12) | **751 (<0.001, -0.529)** |
|  | *Putamen* | *L* | 9.61 (0.29) | 9.43 (0.15) | **885 (<0.001, -0.445)** |
|  |  | *R* | 9.63 (0.27) | 9.40 (0.14) | **791 (<0.001, -0.504)** |
|  | *Pallidum* | *L* | 8.41 (1.08) | 8.88 (0.58) | **2126 (0.004, 0.332)** |
|  |  | *R* | 8.59 (0.94) | 9.05 (0.45) | **2300 (<0.001, 0.447)** |
|  | *Thalamus* | *L* | 9.51 (0.14) | 9.50 (1.32) | 1502 (0.701, -0.059) |
|  |  | *R* | 9.54 (0.13) | 9.52 (0.12) | 1515 (0.723, -0.051) |
| *Extracellular signal fraction* | *Caudate* | *L* | 0.38 (0.04) | 0.36 (0.01) | **757 (<0.001, -0.526)** |
|  |  | *R* | 0.37 (0.02) | 0.36 (0.01) | **984 (<0.001, -0.383)** |
|  | *Putamen* | *L* | 0.36 (0.03) | 0.34 (0.02) | **1048 (0.003, -0.343)** |
|  |  | *R* | 0.34 (0.03) | 0.32 (0.01) | **892 (<0.001, -0.441)** |
|  | *Pallidum* | *L* | 0.36 (0.04) | 0.33 (0.02) | **910 (<0.001, -0.430)** |
|  |  | *R* | 0.35 (0.04) | 0.32 (0.02) | **932 (<0.001, -0.416)** |
|  | *Thalamus* | *L* | 0.34 (0.02) | 0.33 (0.01) | 1294 (0.130, -0.189) |
|  |  | *R* | 0.33 (0.01) | 0.32 (0.01) | 1636 (0.853, 0.025) |
| *Extracellular diffusivity* | *Caudate* | *L* | 1.47 (0.28) | 1.40 (0.11) | 1420 (0.418, -0.110) |
|  |  | *R* | 1.51 (0.25) | 1.44 (0.13) | 1482 (0.633, -0.071) |
|  | *Putamen* | *L* | 1.47 (0.17) | 1.36 (0.11) | **902 (<0.001, -0.435)** |
|  |  | *R* | 1.62 (0.20) | 1.49 (0.11) | **1002 (0.001, -0.372)** |
|  | *Pallidum* | *L* | 1.62 (0.20) | 1.48 (0.12) | **842 (<0.001, -0.472)** |
|  |  | *R* | 1.76 (0.23) | 1.59 (0.14) | **809 (<0.001, -0.493)** |
|  | *Thalamus* | *L* | 1.47 (0.07) | 1.44 (0.07) | 1411 (0.394, -0.116) |
|  |  | *R* | 0.31 (0.03) | 0.31 (0.03) | 1377 (0.305, -0.137) |
| *Apparent neurite density* | *Caudate* | *L* | 0.18 (0.04) | 0.17 (0.02) | 1514 (0.723, -0.051) |
|  |  | *R* | 0.18 (0.05) | 0.16 (0.02) | 1317 (0.167, -0.175) |
|  | *Putamen* | *L* | 0.24 (0.04) | 0.23 (0.03) | 1374 (0.303, -0.139) |
|  |  | *R* | 0.23 (0.04) | 0.23 (0.03) | 1411 (0.394, -0.116) |
|  | *Pallidum* | *L* | 0.41 (0.05) | 0.42 (0.03) | 1553 (0.854, -0.027) |
|  |  | *R* | 0.43 (0.05) | 0.43 (0.03) | 1473 (0.605, -0.078) |
|  | *Thalamus* | *L* | 0.31 (0.03) | 0.31 (0.03) | 1565 (0.868, -0.019) |
|  |  | *R* | 0.31 (0.03) | 0.31 (0.03) | 1459 (0.554, -0.086) |
| *Fractional anisotropy* | *Caudate* | *L* | 0.18 (0.04) | 0.16 (0.02) | **993 (0.001, -0.378)** |
|  |  | *R* | 0.20 (0.07) | 0.16 (0.02) | **889 (<0.001, -0.443)** |
|  | *Putamen* | *L* | 0.19 (0.04) | 0.17 (0.02) | **873 (<0.001, -0.453)** |
|  |  | *R* | 0.20 (0.04) | 0.17 (0.02) | **710 (<0.001, -0.555)** |
|  | *Pallidum* | *L* | 0.28 (0.04) | 0.26 (0.03) | **1115 (0.010, -0.301)** |
|  |  | *R* | 0.28 (0.05) | 0.26 (0.02) | **995.5 (<0.001, -0.401)** |
|  | *Thalamus* | *L* | 0.33 (0.02) | 0.33 (0.02) | 1808 (0.319, 0.133) |
|  |  | *R* | 0.33 (0.02) | 0.33 (0.02) | 1532 (0.776, -0.040) |
| *Mean diffusivity* | *Caudate* | *L* | 6.50<sup>a</sup> (0.41<sup>a</sup>) | 6.23<sup>a</sup> (0.13<sup>a</sup>) | **777.5 (<0.001, -0.513)** |
|  |  | *R* | 6.44<sup>a</sup> (0.36<sup>a</sup>) | 6.20<sup>a</sup> (0.12<sup>a</sup>) | **760.5 (<0.001, -0.523)** |
|  | *Putamen* | *L* | 6.22<sup>a</sup> (0.34<sup>a</sup>) | 5.95<sup>a</sup> (0.15<sup>a</sup>) | **653.5 (<0.001, -0.591)** |
|  |  | *R* | 6.20<sup>a</sup> (0.34<sup>a</sup>) | 5.91<sup>a</sup> (0.13<sup>a</sup>) | **624.5 (<0.001, -0.609)** |
|  | *Pallidum* | *L* | 5.54<sup>a</sup> (0.36<sup>a</sup>) | 5.51<sup>a</sup> (0.23<sup>a</sup>) | 1557.5 (0.854, -0.024) |
|  |  | *R* | 5.52<sup>a</sup> (0.30<sup>a</sup>) | 5.49<sup>a</sup> (0.18<sup>a</sup>) | 1437 (0.474, -0.100) |
|  | *Thalamus* | *L* | 5.98<sup>a</sup> (0.14<sup>a</sup>) | 5.96<sup>a</sup> (0.11<sup>a</sup>) | 1509 (0.721, -0.055) |
|  |  | *R* | 6.01<sup>a</sup> (0.13<sup>a</sup>) | 6.00<sup>a</sup> (0.11<sup>a</sup>) | 1566.5 (0.868, -0.018) |

**Abbreviations:** **FDR:** False Discovery Rate; **HC:** Healthy Controls; **HD:** Huntington's Disease; **L:** Left Hemisphere; **R:** Right Hemisphere; **SD:** Standard Deviation
<sup>a</sup>: Multiplied by $10^{-4}$

The SANDI measures revealed between-group differences in apparent soma density, apparent soma size, and extracellular signal fraction with moderate ES in the BG (*r_rb_* = 0.32-0.53) (Fig. 2). HD individuals exhibited reduced apparent soma density (Fig. 2A) across all BG ROIs. Apparent soma size was elevated in bilateral caudate and putamen but reduced in the pallidum (Fig. 2B). Elevated extracellular signal fraction was observed in all BG regions (Fig. 2C). Extracellular diffusivity was higher in the putamen and pallidum but not in the caudate (Fig. 2D). No differences were observed for apparent neurite density f_in_ in any of the ROIs (*FDR-p* ≥ 0.167, absolute *r_rb_* < 0.2).

Increased FA and MD (*FDR-p* < 0.01) with moderate ES (*r_rb_* = 0.30-0.61) were observed in the BG (Fig. 2E-F) with the exception of MD in the left pallidum (*FDR-p* = 0.854).

No differences were found in the thalami for any of the microstructural metrics (*rrb* = 0.02-0.19).

### Correlations between BG microstructure and motor performance in HD

#### Motor measures

PCA extracted one principal component that explained 64% of the Q-Motor data with high loadings (> 0.5 or <-0.5) from all variables (Supplementary Table 3). Spearman’s rho correlation between Q-motor component scores and the disease burden CAP_100_ score revealed a positive correlation (*ρ* = 0.61, *p* = 0.002, *N* = 24), i.e., higher disease burden was associated with higher scores in the Q-Motor component reflecting slower and less accurate motor performance (Supplementary Figure 1).

Spearman’s rho correlations between Q-motor component scores and SANDI microstructural measures from all ROIs and scatter plots are displayed in Figure 3. Figure 4 displays correlations and scatter plots of the Q-Motor component with DTI and volumetric indices. Overall, negative correlations were observed between principal Q-Motor component scores and apparent soma density in the BG, apparent soma size in the pallidum, and volumetric measurements in all ROIs, indicating that lower apparent soma density and volumes were associated with impaired motor performance, reflected in higher scores in the Q-Motor component. Conversely, positive correlations were present between Q-Motor component scores and apparent soma size, extracellular signal fraction, and extracellular diffusivity in the BG, i.e. larger apparent soma size and extracellular signal were associated with impaired motor performance (Figure 3).

**Figure 3.**
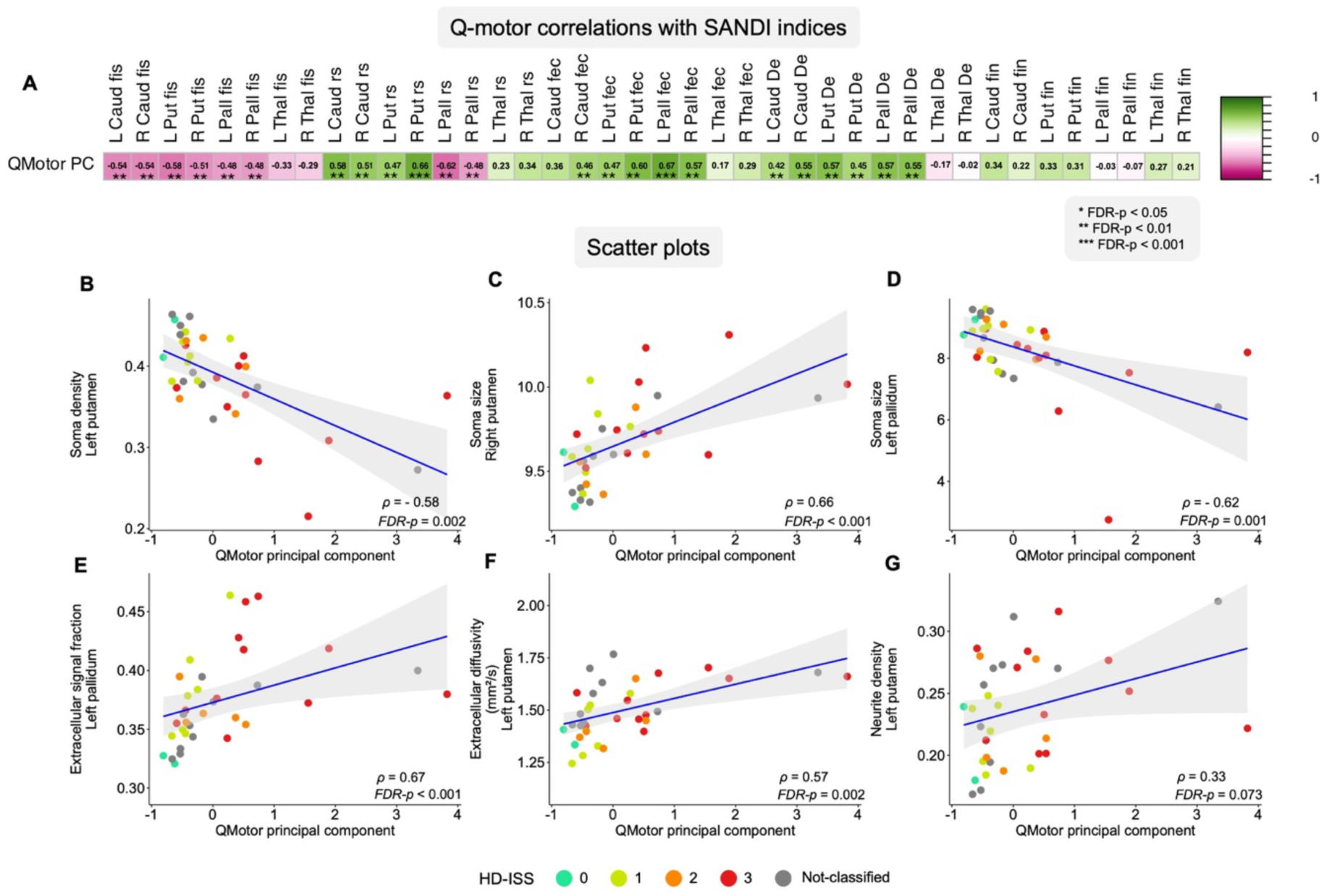
**A)** Correlation matrix and **B-G)** selected scatter plots illustrating Spearman’s rho correlations between SANDI measures and the Q-Motor principal component. **A)** Each cell represents the Spearman’s rho correlation strength, with pink indicating negative and green positive correlations. **B-G)** Each plot includes a best-fit least squares linear regression line with standard error indicated by the grey shaded area, along with the Spearman’s rho (*ρ*) and the corresponding FDR-p value. Regression lines are included for visualisation only and do not reflect variance explained (R²) or imply linear model fit. Scatter dot colours represent participants’ HD-ISS stage. Unclassified refers to those participants who could not be classified due to having CAG 36-40 or incomplete clinical data. **Abbreviations: D_e_**: Extracellular diffusivity; **f_ec_**: Extracellular signal fraction; **f_in_**: Neurite density signal fraction; **f_is_**: Soma density signal fraction; **PC**: Principal component; **r_s_**: Soma radius; **TFC**: Total functional capacity.

**Figure 4.**
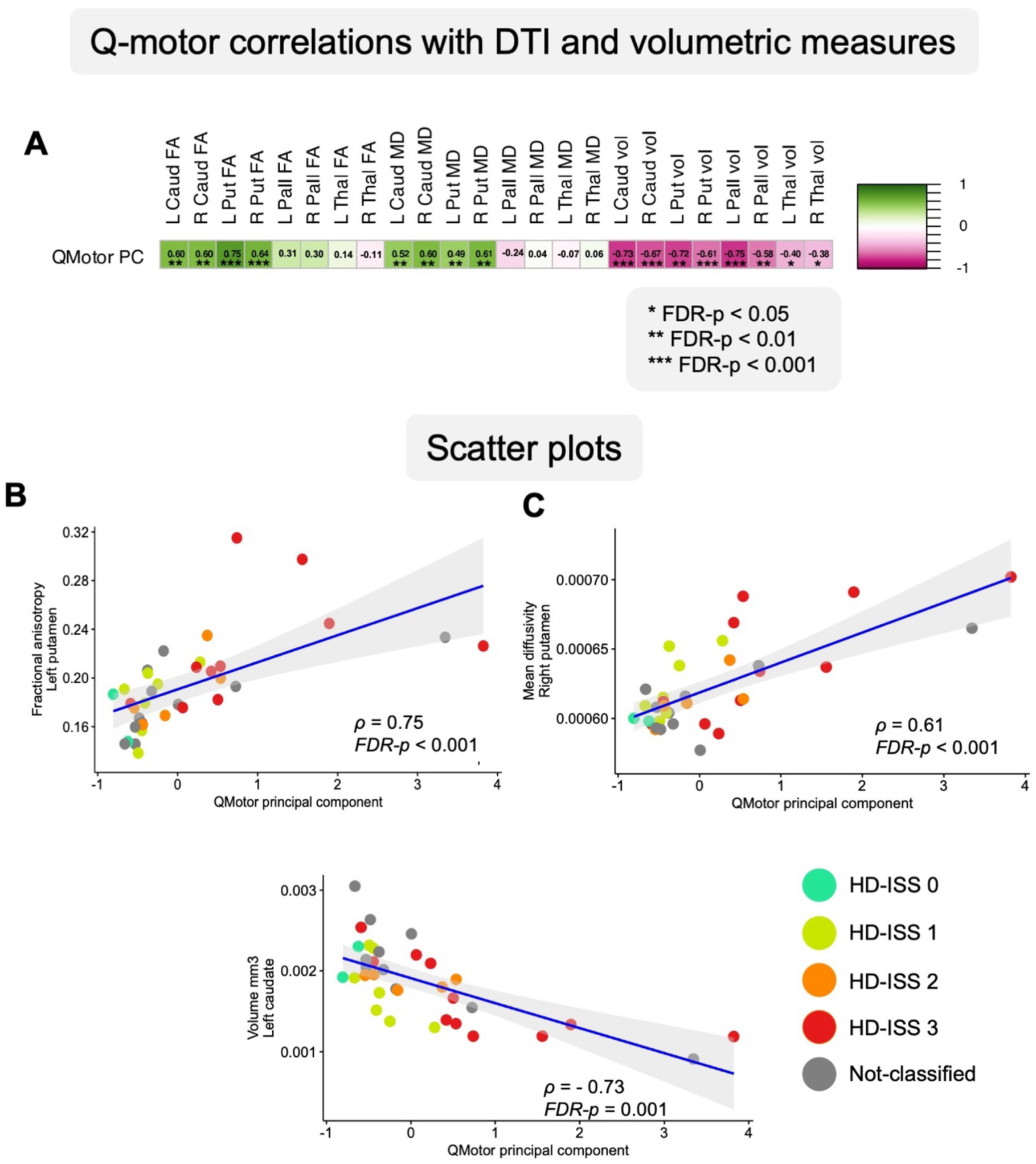
**A)** Correlation matrix and **B-D)** selected scatter plots illustrating Spearman’s rho correlations between DTI, volumetric measures and the Q-Motor principal component. **A)** Each cell represents the Spearman’s rho correlation strength, with pink indicating negative and green positive correlations. **B-D)** Each plot includes a best-fit least squares linear regression line with standard error indicated by the grey shaded area, along with the Spearman’s rho (*ρ*) and the corresponding FDR-p value. Regression lines are included for visualisation only and do not reflect variance explained (R²) or imply linear model fit. Scatter dot colours represent participants’ HD-ISS stage. Unclassified refers to those participants who could not be classified due to having CAG 36-40 or incomplete clinical data. **Abbreviations: FA**: Fractional anisotropy; **MD**: Mean diffusivity.

### Microstructural predictors of BG volumes

Hierarchical linear regression analyses of the HC data (Table 4, Figure 5) revealed that 17% of volumetric variation in the left caudate was accounted for by age and apparent soma density while 11% of variation in the right caudate was explained by age and extracellular diffusivity (D_e_). Age accounted for 25% of volumetric differences in the left putamen, and together with D_e_ for 29% of differences in right putamen. Age alone explained 17% of left and 11% of right thalamic volume variation. No significant regression models were present for bilateral globus pallidus.

**Figure 5.**
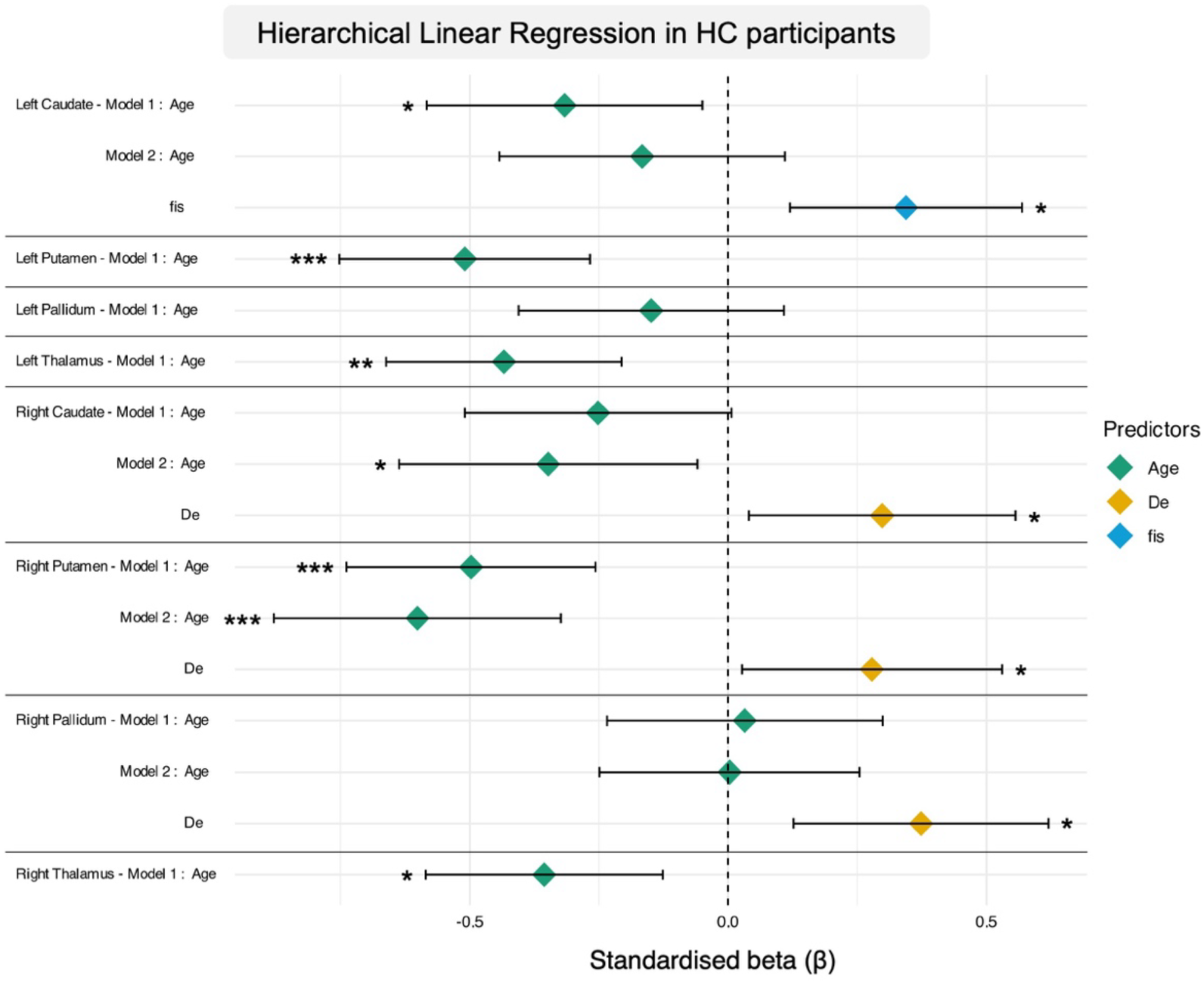
Standardised beta coefficients of SANDI microstructural metrics predicting volume (normalised for intracranial volume) in regions of interest in the healthy control group. Data were modelled by firstly accounting for age, followed by the step-wise inclusion of all SANDI indices. The figure displays the predictor variables included in the final regression models for each region of interest. **Abbreviations: De**: Extracellular diffusivity; **fis**: Soma density signal fraction; **rs**: Soma radius; **TFC:** Total functional capacity. * *p* < 0.05; ** *p* < 0.01; *** *p* < 0.001.

**Table 4.** Hierarchical Linear Regression Predicting Normalised Volumes from SANDI Microstructural Metrics, Controlling for Age in the Healthy Control Participants.

|  |  |  |  |  |  |  |  |  |  |
| --- | --- | --- | --- | --- | --- | --- | --- | --- | --- |
| **ROI** | **Model** | **Predictor(s)** | ***Adjusted R<sup>2</sup>*** | ***ΔR<sup>2</sup>*** | ***F-value (p-value)*** | ***ΔF-value*** | ***β*** | ***t-value*** | ***p-value*** |
| Left Caudate | 1 | Age | 0.084 | 0.100 | **6.115**<br>**(0.017)** |  | -0.316 | -2.473 | **0.031** |
|  | 2 | Age | 0.167 | 0.096 | **6.599**<br>**(0.003)** | 6.474 | -0.166 | -1.226 | 0.297 |
|  |  | f<sub>is</sub> |  |  |  |  | 0.345 | 2.544 | **0.026** |
| Left Putamen | 1 | Age | 0.247 | 0.260 | **19.328**<br>**(<0.001)** |  | -0.510 | - 4.396 | **<0.001** |
| Left Pallidum | 1 | Age | 0.004 | 0.022 | 1.247<br>(0.269) |  | -0.149 | -1.117 | 0.341 |
| Left Thalamus | 1 | Age | 0.174 | 0.188 | **12.761**<br>**(0.001)** |  | -0.434 | -3.572 | **0.003** |
| Right Caudate | 1 | Age | 0.046 | 0.063 | 3.715<br>(0.059) |  | -0.252 | -1.927 | 0.087 |
|  | 2 | Age | 0.111 | 0.080 | **4.508**<br>**(0.015)** | 5.029 | -0.348 | -2.616 | **0.023** |
|  |  | D<sub>e</sub> |  |  |  |  | 0.299 | 2.243 | **0.048** |
| Right Puta-<br>men | 1 | Age | 0.234 | 0.248 | **18.109**<br>**(<0.001)** |  | -0.498 | -4.255 | **<0.001** |
|  | 2 | Age | 0.289 | 0.067 | **12.389**<br>**(<0.001)** | 5.265 | -0.602 | -4.954 | **<0.001** |
|  |  | De |  |  |  |  | 0.279 | 2.295 | **0.045** |
| Right Pal-<br>lidum | 1 | Age | -0.017 | 0.001 | .058 (0.811) |  | 0.032 | 0.241 | 0.847 |
|  | 2 | Age | 0.108 | 0.139 | **4.392**<br>**(0.017)** | 8.718 | 0.003 | 0.025 | 0.980 |
|  |  | De |  |  |  |  | 0.374 | 2.953 | **0.011** |
| Right Thala-<br>mus | 1 | Age | 0.111 | 0.127 | **7.966**<br>**(0.007)** |  | -0.356 | -2.822 | **0.015** |
| **Model 1:** Volume(ROI)=$\beta_0 + \beta_1 \text{*(Age)} + \epsilon$; **Model 2-n:** Volume (ROI)=$\beta_0 + \beta_1 \text{*(Age)} + \beta_{2-n} \text{*(f}_{is}, r_s, f_{ec}, D_e, f_{in}) + \epsilon$; Predictors in Model 1 were entered simultaneously; predictors in Models 2-n were entered in a stepwise fashion using an iterative forward selection and backward elimination method to maximise the adjusted R<sup>2</sup> while keeping only the most significant predictors.<br>**Abbreviations:** **De:** Extracellular diffusivity; **fec:** Extracellular signal fraction; **fin:** Neurite density signal fraction; **fis:** Soma density signal fraction; **rs:** Soma size; **SANDI:** Soma And Neurite Density Imaging; **TFC:** Total Functional Capacity<br>Significant F-values and t-values are highlighted in **bold** |  |  |  |  |  |  |  |  |  |

In contrast, regression analyses for the HD data (Table 5, Figure 6) showed that 60% of volume variation in the left caudate and 51% of variation in the right caudate were accounted for by age and apparent soma density and size. Similarly, 57% of variation in the right putamen volume were explained by age and apparent soma size, while 63% of volume differences in the left putamen were explained by age, apparent soma size as well as extracellular signal and diffusivity. Comparable to the HC results, age alone accounted for 30% of volume variation in the left and for 35% in the right thalamus while no age effects were present for bilateral globus pallidus. However, in HD individuals 42% of volume variation in the left pallidum was explained by apparent soma size and extracellular signal and 27% of volume variation in the right pallidum by extracellular signal fraction only. No significant contributions of TFC were present.

**Figure 6.**
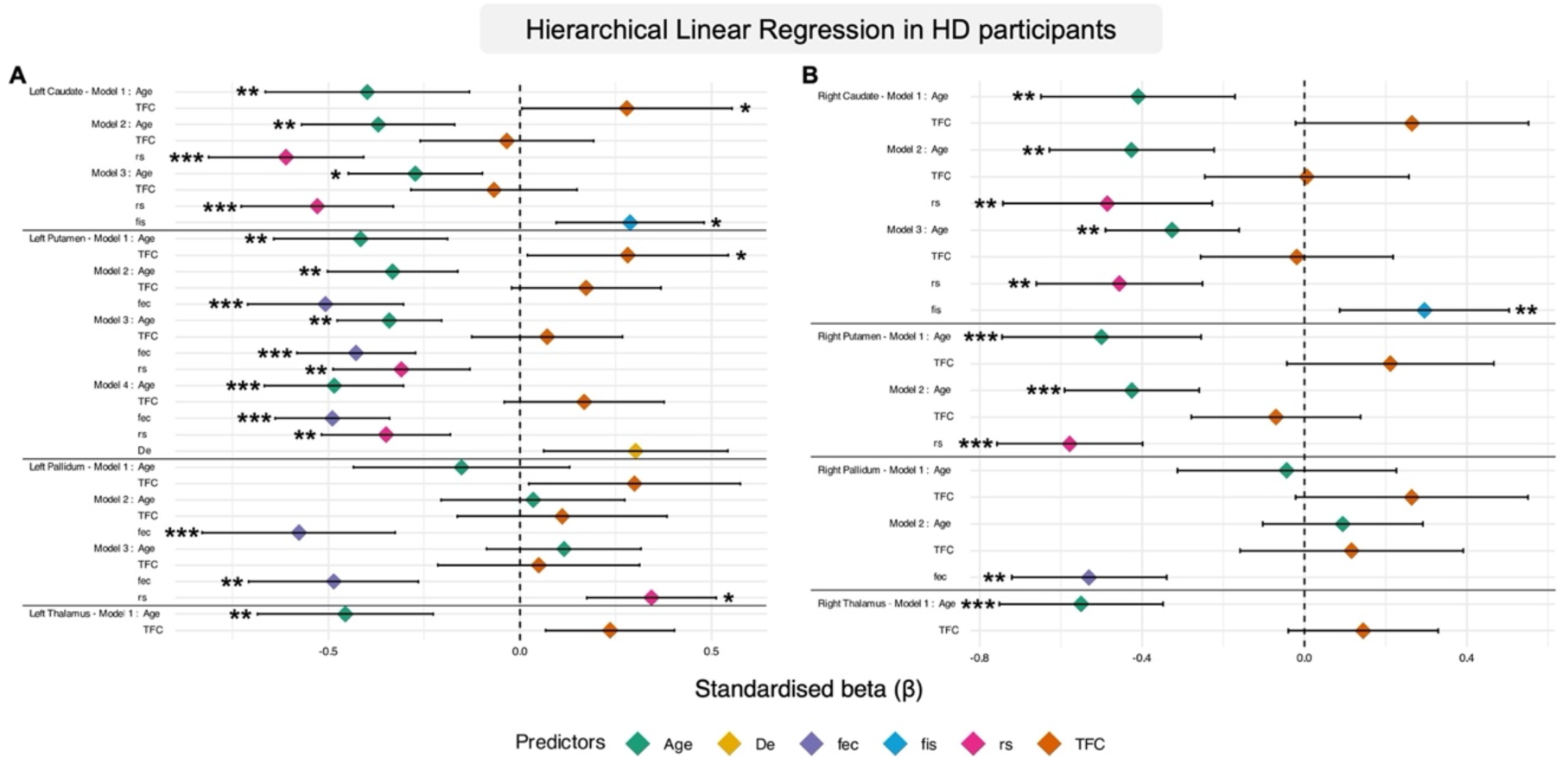
Standardised beta coefficients of SANDI microstructural metrics predicting volume (normalised for intracranial volume) in A) left and B) right hemisphere regions of interest in individuals with Huntington’s disease. Data were modelled by firstly accounting for age and Total Functional Capacity scores simultaneously, followed by the step-wise inclusion of all SANDI indices. The figure displays the predictor variables included in the final regression models for each region of interest. **Abbreviations: De**: Extracellular diffusivity; **fec**: Extracellular signal fraction; **fis**: Soma density signal fraction; **rs**: Soma radius. * *p* < 0.05; ** *p* < 0.01; *** *p* < 0.001.

**Table 5.**
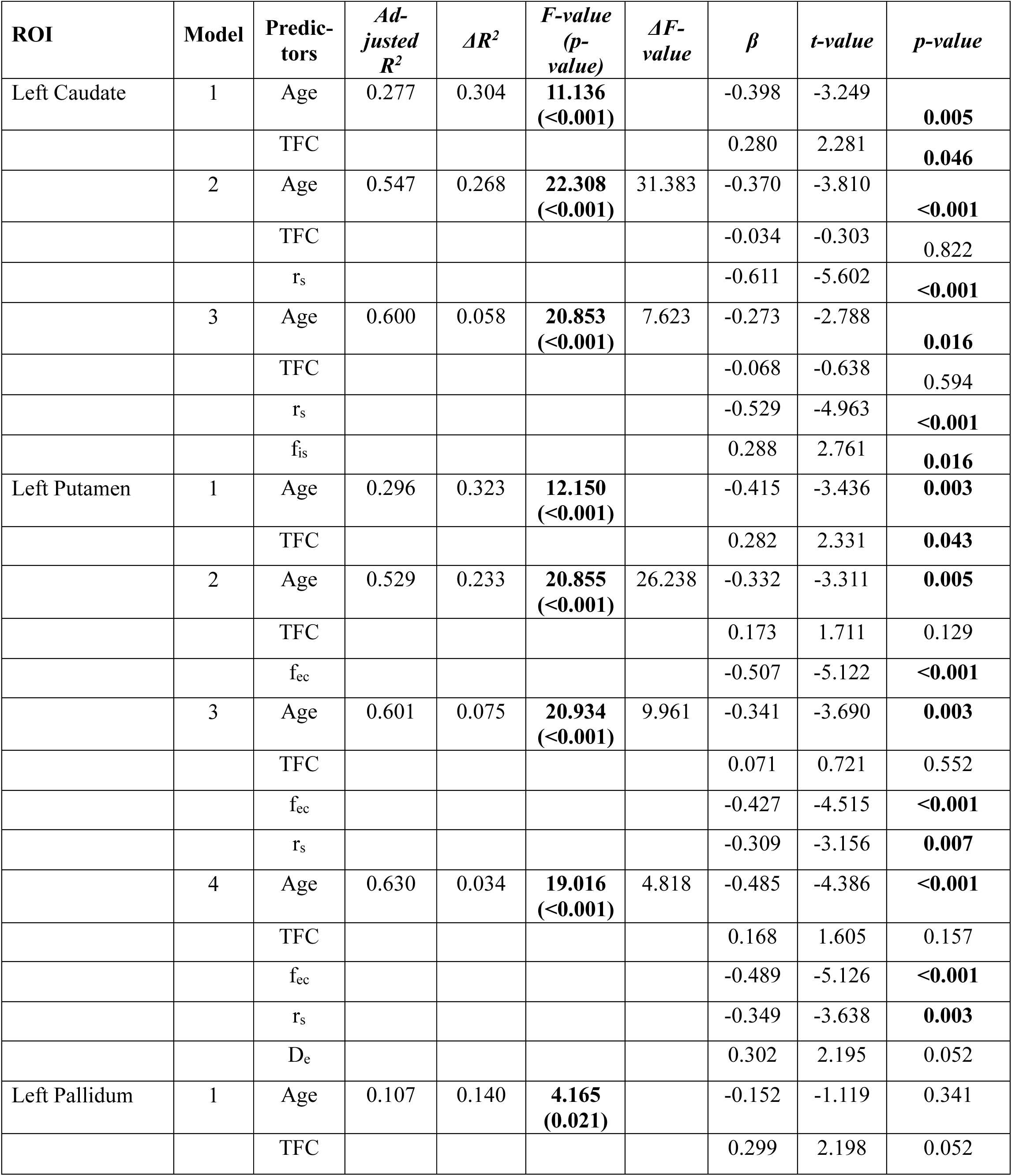
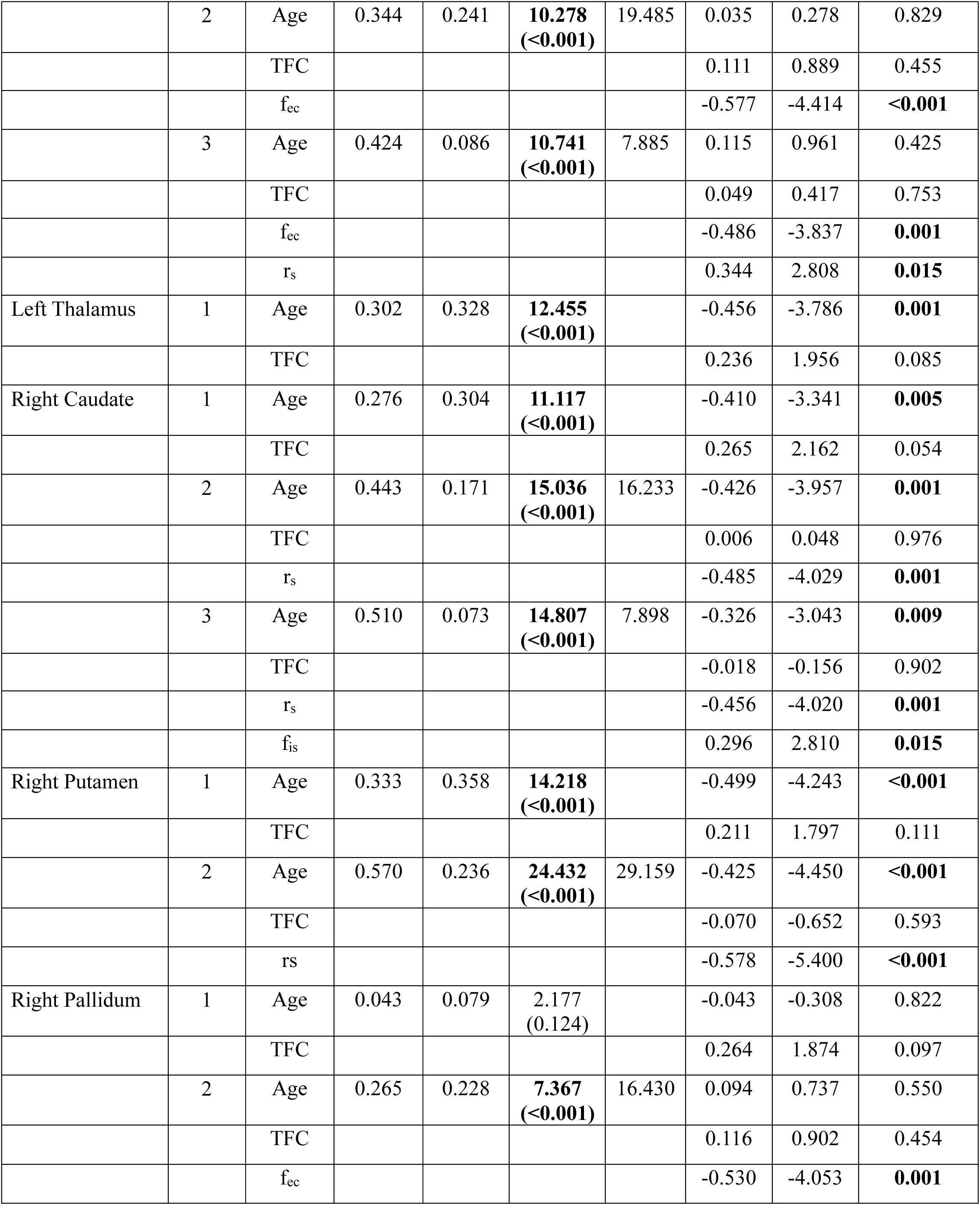
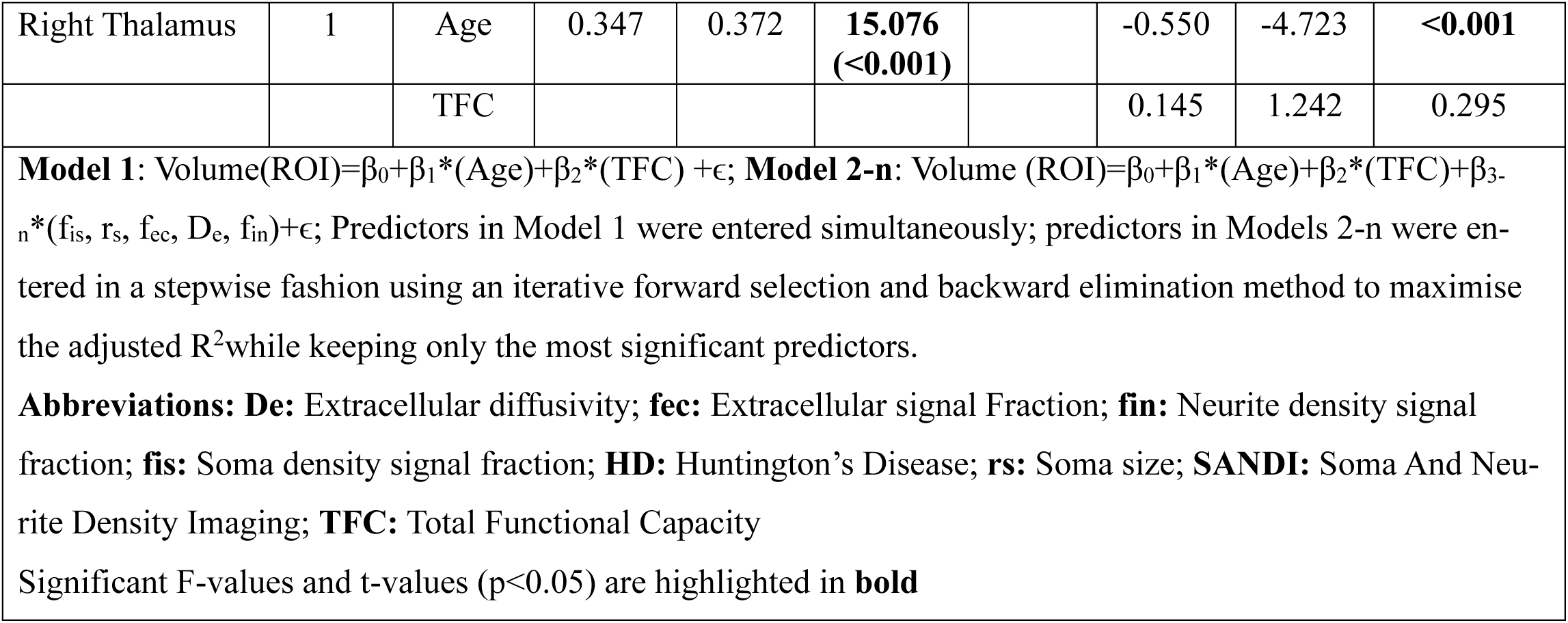
Hierarchical Linear Regression Models Predicting Normalised Volumes in each Region of Interest from SANDI Microstructural Metrics, Controlling for Age and TFC in HD Participants.

### Correlation of disease burden with microstructural and volumetric measures

The CAP_100_ score as an index of disease progression was correlated with brain measurements to explore the relationship between disease burden and microstructural and volumetric differences (Figure 7). Figure 7A summarises correlation coefficient strengths and levels of significance. Figures 7B-J display scatter plots of significant correlations between the CAP_100_ and microstructural and volumetric measures. Increased CAP_100_ was negatively correlated with apparent soma density, and volume size in bilateral caudate and putamen, as well as with pallidal apparent soma size. Positive correlations were observed between CAP_100_ and extracellular diffusivity and signal fraction, apparent soma size in caudate, putamen, and thalamus, apparent neurite density in putamen, FA in the BG, and MD in caudate and putamen.

**Figure 7.**
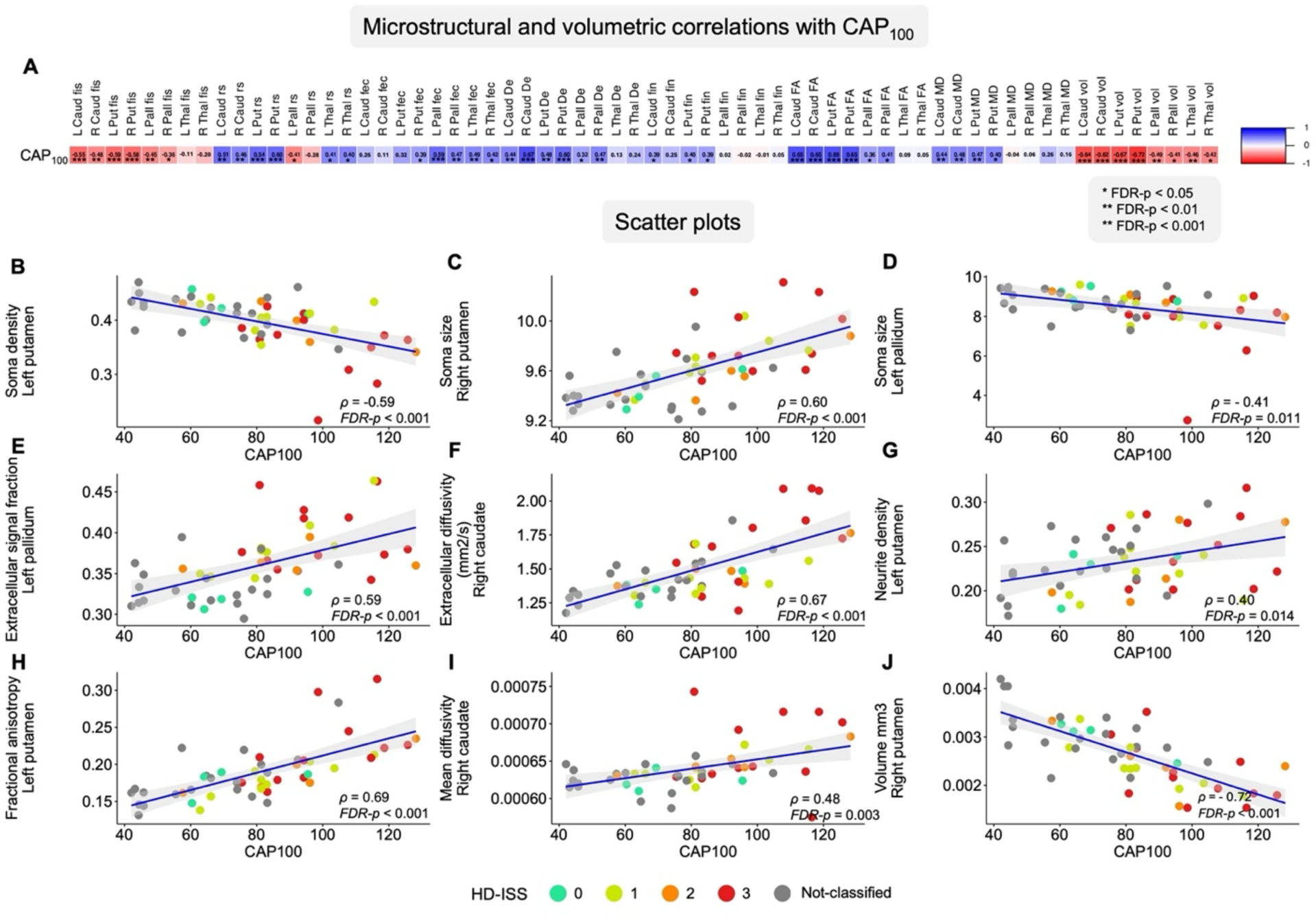
**A)** Correlation matrix and **B-J)** selected scatter plots illustrating Spearman’s rho correlations between SANDI, DTI, and volumetric measures with CAP_100_. Each scatter plot includes a best-fit least squares linear regression line with standard error indicated by the grey shaded area, along with the Spearman’s rho (*ρ*) and the corresponding FDR-p value. Regression lines are included for visualisation only and do not reflect variance explained (R²) or imply linear model fit. Scatter dot colours represent participants’ HD-ISS stage and those who were not classified due to having CAG<40 or incomplete clinical data. **Abbreviations: De**: Extracellular diffusivity; **FA**: Fractional anisotropy; **fec**: Extracellular signal fraction; **fin**: Neurite density signal fraction; **fis**: Soma density signal fraction; **MD**: Mean diffusivity; **rs**: Soma radius; **vol**: Normalised volume.

### Exploratory pairwise comparisons between HD-ISS groups and HC

Exploratory comparisons of apparent soma density, apparent soma size, extracellular signal fraction and extracellular diffusivity, averaged across left and right BG ROIs, were conducted between HD-ISS 0-1 (n = 13), HD-ISS 2-3 (n = 17), and HC (Supplementary Table 1).

Compared with HC, HD-ISS 0-1 showed increased extracellular signal fraction in the BG, larger apparent soma size in the caudate and putamen, and lower apparent soma density alongside larger extracellular diffusivity in the pallidum (*r*_rb_ = -0.255–0.483, *p* ≤ 0.033). Relative to HD-ISS 0-1, those at HD-ISS 2-3 exhibited reduced apparent soma density and increased extracellular diffusivity in the caudate and putamen, as well as smaller apparent soma size in the pallidum (*r*_rb_ = -0.439–0.447, *p* = 0.01–0.031). Compared with HC, the HD-ISS 2-3 group showed differences in the same directions across all metrics in all BG regions. Descriptive statistics and full statistical analysis results are provided in Supplementary Table 4, and effect sizes with 95% confidence intervals are shown in Supplementary Figure 2.

### Cross-correlations between microstructural and volumetric measures

Cross-correlation matrices between SANDI microstructural indices, DTI metrics, and normalised volumes are shown in Supplementary Figure 3 for the full sample, and separately for HC and HD groups. Across all participants, moderate to strong correlations were observed between conceptually related diffusion-derived measures (e.g. mean diffusivity with extracellular diffusivity, fractional anisotropy with neurite-related indices), indicating partially shared variance as expected. In the HC group, SANDI metrics reflecting apparent soma properties (soma radius and soma fraction) showed distinct correlation patterns: soma and neurite fraction were strongly inversely correlated (*ρ* = -0.92) but neither correlated with soma radius. All SANDI measures correlated weakly with FA and volume and moderately with MD. Group-specific matrices revealed broadly similar correlation structures in HC and HD participants. However, several associations, particularly between SANDI indices and normalised volume, were stronger in the HD group, consistent with disease-related coupling between microstructural alterations and macroscopic atrophy.

## Discussion

This study is the first to investigate HD-related microstructural differences in the BG using SANDI, a diffusion MRI technique designed to probe grey matter microstructure. The objective of the study was to explore SANDI indices as potential non-invasive *in vivo* markers of HD neuropathology that may offer greater specificity than volumetric measurements. Apparent soma density and apparent soma size were sensitive to HD-related differences in the BG and together with age, accounted for up to 63% of striatal atrophy. SANDI indices also correlated with motor impairments and CAP_100_ disease burden. Together, these findings highlight the potential of SANDI metrics as imaging biomarkers of disease progression and as surrogate outcome measures for assessing the neural effects of emerging disease-modifying therapies in HD and other neurodegenerative diseases.

### Microstructural and Volumetric Alterations in HD

Well-established patterns of marked volume loss accompanied by increases in FA and MD in the BG were replicated in HD compared with healthy controls. FA increases in the caudate and putamen are thought to reflect the selective degeneration of MSN connections.^20,84^ No microstructural differences and only trends for volumetric reduction were observed in the thalami. This pattern of macro- and microstructural differences in the BG aligns with previous reports in premanifest and early manifest HD stages.^31,85^

Importantly, SANDI revealed novel HD-related microstructural differences in the BG. Compared with healthy controls, those with HD showed reduced apparent soma density together with increased extracellular signal fraction and diffusivity across the caudate, putamen, and pallidum, but not the thalami. Apparent soma size was also increased in the caudate and putamen, whereas it was reduced in the globus pallidus.

HD-related reductions in apparent soma density in the BG are consistent with the loss of striatal MSN, the histopathological hallmark of HD,^22,23^ and with downstream degeneration of pallidal neurons by the loss of striatal projections. Changes in apparent soma size may reflect alterations in neural and glial cell proportions and/or morphology, including astrocyte and microglia swelling in response to neurodegeneration^22,24–26^ as well as soma shrinkage preceding neuronal cell death.^86,87^ Thus, increased apparent soma size in the striatum may indicate HD-related reorganisation of cell composition driven by MSN loss and reactive glial cell swelling, whereas smaller apparent soma size in the pallidum may reflect infiltration of smaller glia cells associated with secondary neuronal loss following striatal degeneration.

Exploratory analyses of BG SANDI differences across HD-ISS Stages 0-3 (Supplementary Figure 2) showed that reductions in striatal apparent soma density and increases in extracellular diffusivity were evident in HD-ISS 2-3 but not in HD-ISS 0-1, whereas increases in striatal apparent soma size and extracellular signal fraction were present in both groups. In the pallidum, reductions in apparent soma density and increases in extracellular signal fraction and diffusivity were observed in both HD-ISS groups, while reduced apparent soma size was only present in HD-ISS 2-3. These findings suggest that SANDI indices are sensitive to changes in BG cellular composition both at early (HD-ISS 0-1) and across disease stages (HD-ISS 0-3). However, these preliminary observations require validation in larger, prospective cohorts before SANDI indices can be considered reliable biomarker of disease progression.

With the development of disease-modifying therapies for HD, the need for non-invasive imaging markers that can sensitively capture treatment effects has become increasingly important. Striatal volume remains the gold-standard imaging marker, and DTI has been widely used to assess tissue microstructure in both cross-sectional and longitudinal studies.^2,13–15,88^ An important question therefore concerns how SANDI indices compare with these established imaging markers and the extent to which they share variance.

In this study we observed moderate effect sizes for group differences in striatal SANDI indices, comparable to those for volume, FA, and MD. ES for apparent soma size overlapped with those for volume and MD, while apparent soma density showed slightly lower but broadly similar ES to FA.

Multicollinearity between microstructural and volumetric imaging measures was assessed using cross-correlations between all imaging metrics (SANDI, DTI, volume) averaged across ROIs (see Supplementary Figure 2). In HCs, apparent soma and neurite fractions were strongly inversely correlated but neither correlated with soma radius. SANDI indices showed only weak correlations with FA and volume, but moderate correlations with MD. This pattern suggests that soma density and size parameters capture microstructural features that are distinct from neurite fraction, FA, and volume, but overlap to some extent with MD, an unspecific metric influenced by multiple biological and geometrical tissue properties.^89,90^ In HD participants, correlations between SANDI metrics and volumes were stronger, and soma radius and fraction were inversely correlated, likely reflecting disease-related reorganisation of microstructural properties associated with atrophy.

### Microstructural predictors of basal ganglia atrophy in HD

To explore the relationship between SANDI microstructural and volumetric measures in the BG, hierarchical linear regression analyses were conducted for each ROI and for HD and HC groups separately, testing age and SANDI indices as predictors of volume. For the HD group, TFC was included as a marker of disease burden. These analyses showed that SANDI indices accounted for a substantial proportion of BG atrophy but not thalamic volume.

Up to 63% of HD-related striatal atrophy was explained by apparent soma density, apparent soma size, and age. In the pallidum, apparent soma size and extracellular signal fraction accounted for 27% of atrophy on the right and 42% on the left, whereas age alone predicted thalamic volume. This pattern in the thalami mirrored findings in healthy controls, where age was the strongest predictor across all ROIs (except bilateral pallidum) and the only significant predictor in the thalami; diffusivity contributed to right-lateralised BG regions and apparent soma density to the left caudate.

Together, these results demonstrate that SANDI-derived measures of apparent soma density and size capture HD-related differences in striatal grey-matter microstructure *in vivo*. As outlined above, reductions in apparent soma density and increases in apparent soma size are consistent with the characteristic loss of medium spiny neurons and reactive gliosis in HD and explain a significant proportion of atrophy in the caudate and putamen.

### Associations Between SANDI microstructural indices and Motor Function

The observed correlations between SANDI metrics and motor measures provide novel insights into the functional implications of microstructural alterations in HD. Consistent with the role of the BG in motor initiation and coordination,^91^ the present study showed that reduced apparent soma density in the striatum and reduced apparent soma size in the pallidum were associated with poorer Q-Motor performance, which itself was linked to greater disease burden. This was reflected in increased IOI and ACU during speeded tapping tasks, and in impaired performance on paced tapping tasks. Similarly, increased apparent soma size in the striatum, together with elevated extracellular signal and diffusivity across all BG regions, as well as striatal FA and MD, were associated with motor impairments. These findings indicate that microstructural changes related to BG neurodegeneration and glial reactivity are directly linked to subtle motor deficits in HD.

### Limitations

HD-ISS classification was possible for only 30 of the 56 HD participants (54%) due to the retrospective pooling of MRI datasets collected before HD-ISS information was incorporated into the Enroll-HD database, missing clinical data, and HD-ISS model exclusions for individuals with CAG repeat lengths between 36-39. Of the 30 classified participants, 4 were assigned to i Stage 0, 9 to Stage 1, 5 ito Stage 2, and 12 to Stage 3; none were classified as Stage 4. Because of the small sample sizes, HD-ISS comparisons were conducted on combined groups (HD-ISS 0-1 versus HD-ISS 2-3). These combined groups showed some overlap in imaging and behavioural features (Figures 3, 4, and 7) and therefore do not represent discrete disease stages. Consequently, these exploratory findings should be interpreted with caution and require replication in larger, prospective cohorts before SANDI metrics can be considered markers of disease progression.

In addition, several methodological considerations regarding HD-ISS classification should be noted. Classification was obtained either from Enroll-HD or calculated using the online HD-ISS tool. For participants labelled “<2” in Enroll-HD, Stages 0 and 1 were assigned based on z-scores for striatal volumes derived from the FreeSurfer v6 cross-sectional pipeline. However, HD-ISS classification and online calculator were developed using volume cut-offs generated from the longitudinal FreeSurfer v6 pipeline. Consequently, applying cross-sectional data in this study may have introduced classification bias.

Furthermore, hierarchical regression models were fitted separately within HD and HC groups and were not intended to support formal statistical comparisons of regression coefficients across groups. Rather, these analyses were designed to explore within-group associations and to provide a descriptive comparison of patterns of relationships linking microstructural indices to regional atrophy. Hierarchical regression models may also be susceptible to over-fitting, and therefore the reported coefficients may not reflect true out-of-sample R^2^.

The assessment of potential multicollinearity between SANDI-derived metrics was based primarily on the healthy control data, as correlations observed in the HD group likely reflect disease-related shifts in microstructural relationships rather than intrinsic model dependencies. In healthy controls, soma radius did not correlate with apparent soma or neurite fractions, whereas a pronounced inverse relationship was observed between soma and neurite fractions, consistent with the biophysical constraints of the model. Importantly, correlations between SANDI metrics and conventional DTI measures (FA) and regional volume were generally weak, indicating that apparent soma estimates capture distinct aspects of grey matter microstructure rather than redundant information.

Moderate correlations with mean diffusivity were observed and are expected, given that MD is a non-specific metric sensitive to multiple microstructural features and susceptible to partial volume effects associated with tissue loss. Consequently, individual regression coefficients should not be interpreted in isolation as fully independent biological effects. As with all model-based approaches applied to correlated imaging parameters, some degree of shared variance and parameter coupling is unavoidable. As we cannot rule out some degeneracy between apparent soma density f_is_ and apparent soma size r_s_, 𝑟_𝑠_ should not be interpreted as absolute histological soma radii, and inferences should instead focus on relative group differences and spatial patterns. Similarly, HD-related microstructural changes (e.g., changes to membrane permeability) could affect model parameter fidelity, and thus 𝑓_𝑖𝑠_and 𝑟_𝑠_should be treated as MRI-derived effective indices rather than direct quantitative measures of neuron loss or glial hypertrophy.

However, the correlation structure observed in healthy controls suggests that multicollinearity is unlikely to critically undermine model stability or interpretability in the present analyses. Importantly, all comparisons were made under an identical acquisition protocol and fitting pipeline, meaning that group-level contrasts remain informative even if absolute parameter values are biased. The primary conclusions of the present study therefore do not rely on the precise magnitude of individual coefficients but on the consistent pattern and direction of associations across regions and measures. Together, these factors support the feasibility and initial clinical relevance of SANDI for probing striatal microstructural alterations in Huntington’s disease.

### Clinical Implications and Future Directions

The present study acquired multi-shell (max b-value = 6,000 s/mm^2^) DWI data on a non-clinical ultra-strong gradient (300mT/m) 3T MRI system. Ultra-strong gradients improve the signal-to-noise-ratio at high b-values by enabling shorter echo times (TE), thereby enhancing sensitivity to small water displacement,^39^ and reducing bias from inter-compartmental exchange.^92,93^

However, for clinical translation it is important to demonstrate that HD-related SANDI differences can be detected on standard clinical MRI systems without requiring ultra-strong gradients. Although equivalent data in HD are not yet available, we have shown the feasibility of estimating SANDI metrics from multi-shell DWI acquired on a 3T MRI system with standard gradient strength (67mT/m) (maximum b-value of 6,000 s/mm^2^) in healthy adults ^94^ and people with MS^42,44,94^. Additionally, Zeng et al.,^43^ reported significant differences in SANDI metrics between individuals with ALS and healthy controls using a 3T MRI Prisma system with a maximum b-value of 3,000 s/mm^2^, and two further studies using 3T clinical scanners with standard gradient strength showed detectable microstructural SANDI alterations in MS. Collectively, these findings indicate that SANDI can be implemented on clinical scanners, particularly as commercial systems progress toward stronger gradient capabilities such as Siemens Magnetom Cima.X.

Our findings highlight the potential of SANDI-derived metrics as future markers for tracking disease progression and assessing therapeutic efficacy in HD, as well as in more prevalent neurodegenerative conditions such as Alzheimer’s and Parkinson’s disease. The sensitivity of apparent soma density and size, and extracellular water signal to microstructural changes in HD provides information complementary to volumetric measures, which are currently the most widely used imaging modality in clinical trials. Furthermore, the observed associations between SANDI metrics, motor performance, and disease burden underscore their relevance for evaluating the neural effects of emerging disease-modifying treatments.

Ultimately, the choice of imaging markers depends on scientific or clinical objective: DTI and volumetric measures are informative for documenting differences or tracking longitudinal change, whereas gaining mechanistic insight into tissue microstructure and therapeutic effects requires going beyond DTI and volumes. Advanced models such as SANDI offer the potential to provide more specific and biologically meaningful characterisation of grey matter pathology.

## Conclusion

Our study demonstrates the potential of SANDI for characterising microstructural abnormalities in individuals with HD. These abnormalities align with *ex vivo* neuropathological findings of striatal medium spiny neuron loss and gliosis, account for a substantial proportion of striatal atrophy, and correlate with motor impairment and disease burden. Although conventional MRI lacks the resolution to directly capture histopathology, advanced biophysical models such as SANDI may help bridge this gap by providing biologically interpretable parameters that reflect tissue composition and capture histopathological changes *in vivo*. This approach offers a promising avenue for advancing HD research and improving clinical care.

## Data availability

This research utilised baseline data from the HD-DRUM project that has been endorsed by the Enroll-HD Scientific Oversight Committee (SOC) (14/11/2022). At the end of the HD-DRUM project, the coded study data will be shared and made accessible to the research community *via* the Enroll-HD specific data request process. WAND data are publicly available (https://git.cardiff.ac.uk/cubric/wand).

## Data Availability

This research utilised baseline data from the HD–DRUM project that has been endorsed by the Enroll–HD Scientific Oversight Committee (SOC) (14/11/2022). At the end of the HD–DRUM project, the coded study data will be shared and made accessible to the research community via the Enroll–HD specific data request process. WAND data are publicly available (https://git.cardiff.ac.uk/cubric/wand).

## Acknowledgements

We would like to thank Amy Dangerfield, Allison Cooper and Sonya Foley-Bozorgzad for their help with data collection as well as Derek Jones and John Evans for their advice and support with regards to the implementation of MRI data acquisition protocols. We would like to thank the following clinical and administrative staff at the participating patient identification centres for their help with identifying suitable patients for the study: Eileen Donovan, Kim Munnery, and Jane Davies from the Cardiff HD clinic; Jessica Prado Mota from the Royal Devon University Healthcare NHS Foundation Trust in Exeter; Jenni Burns from the Walton Centre NHS Foundation Trust in Liverpool; Natalie Rosewell, Anya Soonderpershad, and Dr Liz Coulthard from the Bristol Brain Centre; Claire Tilley and Dr Hugh Rickards from the Birmingham and Solihull Mental Health NHS Foundation Trust. In addition, we would like to thank all Public Involvement contributors and the members of the Enroll-HD Scientific Oversight Committee for their input into the study as well as all participants for their generous time commitment to help us conducting this research.

## Supplementary figure legends

**Supplementary Figure 1.**
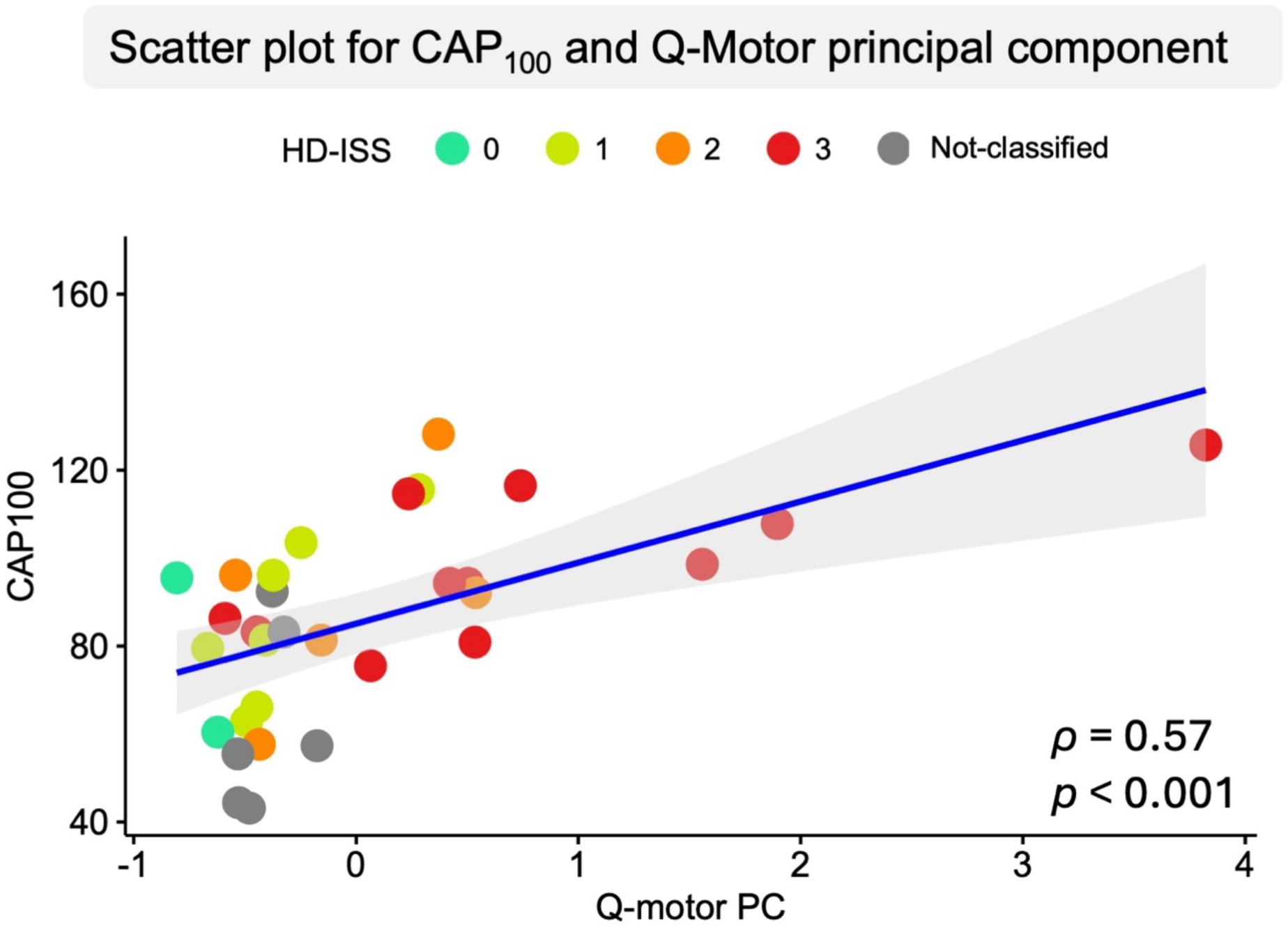
Scatterplot showing positive relationship between the Q-Motor principal component and the disease burden measure (CAP_100_) with the Spearman’s rho (*ρ*) test. Scatter dot colours represent participants’ HD-ISS stage and those who were not classified due to having CAG<40 or incomplete clinical data.

**Supplementary Figure 2.**
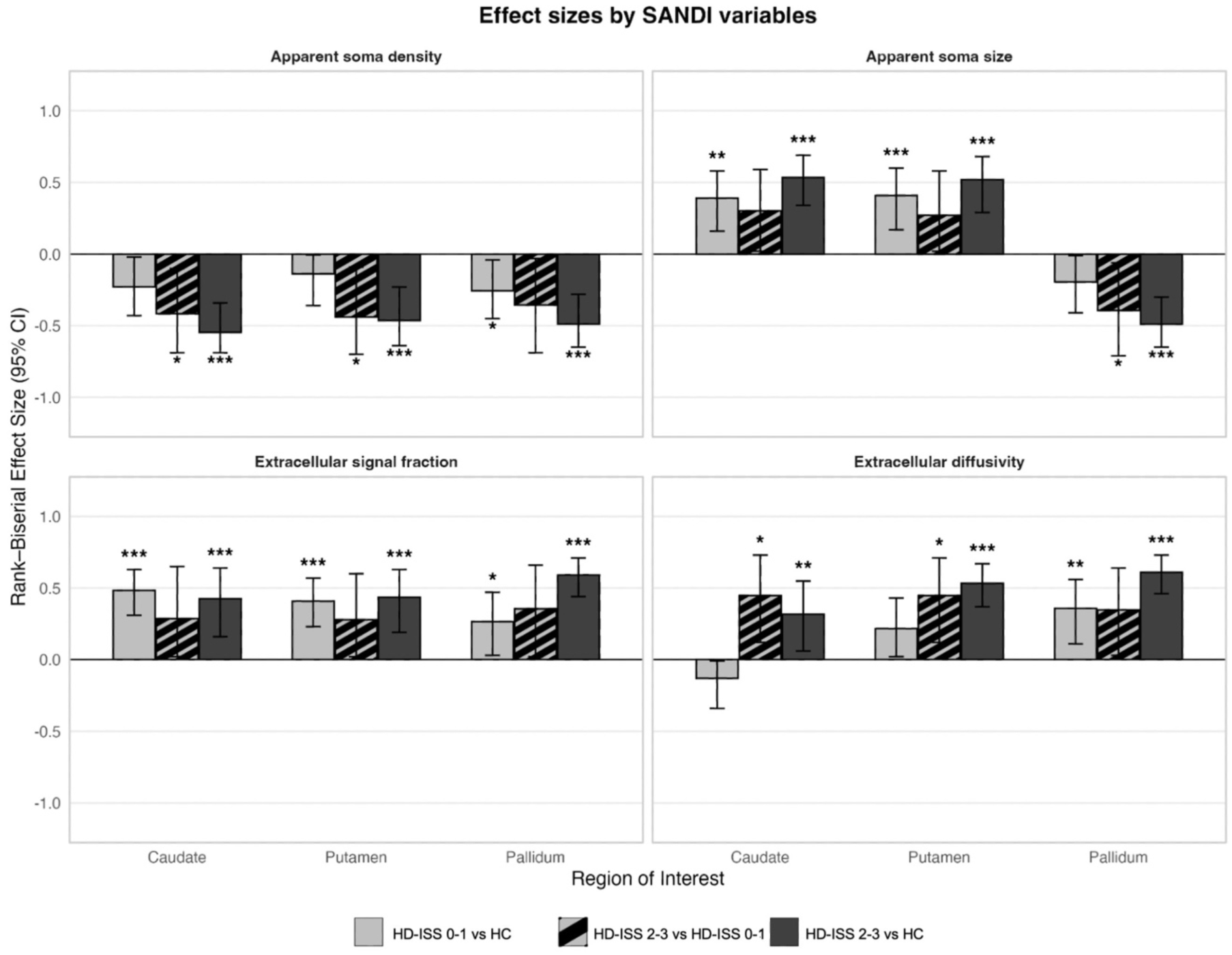
Bar plot showing the effect sizes and 95% confidence intervals for exploratory pairwise comparisons between HD-ISS 0-1 (n = 13), HD-ISS 2-3 (n = 17), and healthy controls (HC). Significant comparisons are marked with * (p < 0.05), ** (p < 0.01), and *** (p < 0.001).

**Supplementary Figure 3.**
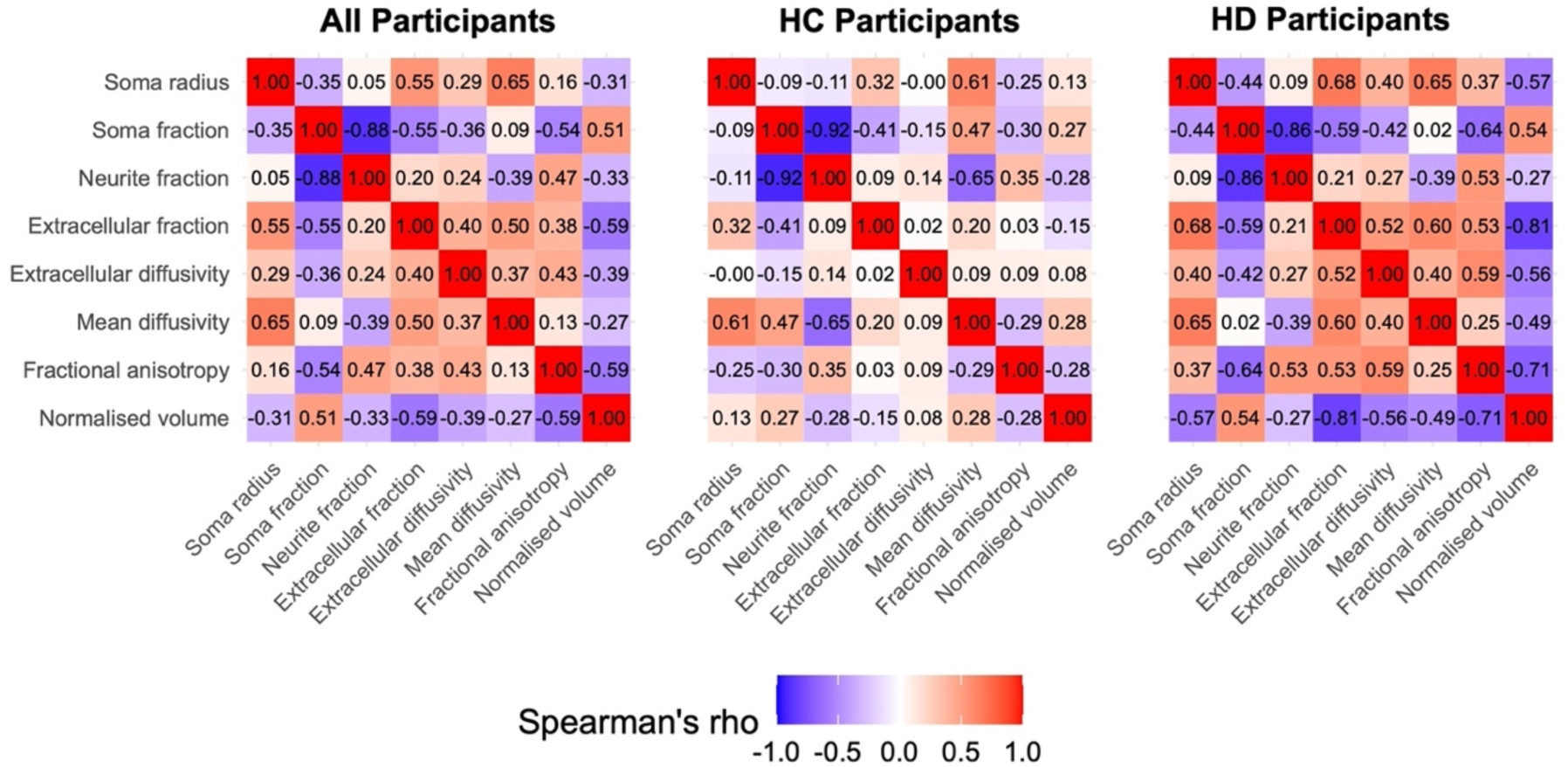
Correlation heatmaps showing the cross-correlation of SANDI, DTI, and volumetric (normalised for intracranial volume) measures, averaged across the caudate, putamen, pallidum, and thalamus. Heatmaps are shown separately for the full sample, healthy controls (HC), and Huntington’s disease (HD) participants. Correlations are expressed as Spearman’s rho coefficients.

**Supplementary Table 1.** Demographic and clinical information per HD-ISS stage.

|  | HD-ISS 0 |  | HD-ISS 1 |  | HD-ISS 2 |  | HD-ISS 3 |  | HD-ISS NA |  |
| --- | --- | --- | --- | --- | --- | --- | --- | --- | --- | --- |
| Measure | N | Mean (SD) | N | Mean (SD) | N | Mean (SD) | N | Mean (SD) | N | Mean (SD) |
| Age | 4 | 37.75 (16.30) | 9 | 47.11 (12.36) | 5 | 48.80 (10.83) | 12 | 49.17 (12.20) | 26 | 42.65 (14.97) |
| Sex, Female (%) | 4 | 3 (75%) | 9 | 5 (56%) | 5 | 1 (20%) | 12 | 3 (25%) | 26 | 13 (50%) |
| Education (years) | 2 | 15.50 (0.71) | 7 | 15.71 (1.38) | 5 | 14.60 (3.71) | 12 | 13.00 (2.17) | 12 | 14.00 (3.13) |
| MOCA | 4 | 25.50 (1.91) | 9 | 28.00 (1.87) | 5 | 26.00 (2.45) | 11 | 23.73 (3.20) | 26 | 27.35 (3.94) |
| TOPF | 4 | 60.25 (5.38) | 9 | 50.22 (14.03) | 5 | 44.20 (14.10) | 12 | 43.25 (10.34) | 26 | 51.92 (13.67) |
| UHDRS-TFC | 4 | 13.00 (0.00) | 9 | 12.78 (0.44) | 5 | 13.00 (0.00) | 12 | 10.92 (1.16) | 24 | 12.12 (1.30) |
| UHDRS-TMS | 4 | 1.50 (3.00) | 9 | 4.67 (4.90) | 5 | 10.20 (1.48) | 12 | 29.33 (20.38) | 18 | 6.22 (11.34) |
| CAG | 4 | 43.00 (2.16) | 9 | 41.89 (1.17) | 5 | 42.00 (1.00) | 12 | 43.67 (3.14) | 20 | 40.40 (2.56) |
| CAP100 | 4 | 72.34 (15.89) | 9 | 85.50 (16.98) | 5 | 91.09 (25.58) | 12 | 99.70 (16.68) | 20 | 65.84 (18.50) |
| SDMT | 2 | 69.00 (5.66) | 7 | 54.29 (13.03) | 5 | 45.60 (8.73) | 12 | 33.83 (12.65) | 12 | 48.00 (12.62) |

**Abbreviations:** **CAG:** Cytosine Adenine Guanine; **CAP:** CAG-Age-Product; **HC:** Healthy controls; **HD:** Huntington's disease; **HD-ISS:** HD-Integrated Staging System; **MOCA:** Montreal Cognitive Assessment; **SD:** Standard deviation; **SDMT:** Symbol Digit Modalities Task; **TFC:** Total Functional Capacity; **TMS:** Total Motor Score; **TOPF:** Test of Premorbid Functioning; **UHDRS:** United Huntington's Disease Rating Scale

**Supplementary Table 2.** Descriptive statistics for motor outcome measures.

| Task<br>*Outcome measure* | HD group |  |
| --- | --- | --- |
|  | N | Mean (SD) |
| **Speeded tapping** |  |  |
| *Left finger; Area under the curve (N-s)* | 38 | 0.13 (0.20) |
| *Right finger; Area under the curve (N-s)* | 38 | 0.07 (0.09) |
| *Left finger; Inter-onset interval (s)* | 38 | 0.26 (0.12) |
| *Right finger; Inter-onset interval (s)* | 38 | 0.23 (0.08) |
| *Left foot; Inter-onset interval (s)* | 37 | 0.37 (0.20) |
| *Right foot; Inter-onset interval (s)* | 38 | 0.35 (0.18) |
| **Metronome tapping; Absolute deviation from pace (s)** |  |  |
| *Left finger; Paced; Fast* | 38 | 0.07 (0.06) |
| *Left finger; Paced; Slow* | 38 | 0.13 (0.14) |
| *Left finger; Unpaced; Fast* | 38 | 0.07 (0.07) |
| *Left finger; Unpaced; Slow* | 38 | 0.15 (0.15) |
| *Right finger; Paced; Fast* | 38 | 0.07 (0.06) |
| *Right finger; Paced; Slow* | 38 | 0.13 (0.12) |
| *Right finger; Unpaced; Fast* | 38 | 0.08 (0.06) |
| *Right finger; Unpaced; Slow* | 38 | 0.16 (0.17) |
| *Left foot; Paced; Fast* | 36 | 0.07 (0.05) |
| *Left foot; Paced; Slow* | 35 | 0.16 (0.13) |
| *Left foot; Unpaced; Fast* | 36 | 0.08 (0.09) |
| *Left foot; Unpaced; Slow* | 35 | 0.17 (0.14) |
| *Right foot; Paced; Fast* | 37 | 0.08 (0.04) |
| *Right foot; Paced; Slow* | 37 | 0.16 (0.12) |
| *Right foot; Unpaced; Fast* | 37 | 0.09 (0.09) |
| *Right foot; Unpaced; Slow* | 37 | 0.16 (0.13) |
| **Pointing (dominant hand)** |  |  |
| *Target frequency over trial (Hz)* | 38 | 2.62 (0.65) |
| **Pointing (dominant) and tapping (non-dominant)** |  |  |
| *Finger tapping area under the curve (coefficient of variation %)* | 38 | 0.76 (0.37) |
| *Finger tapping area under the curve (N-s)* | 38 | 0.31 (0.43) |
| *Finger tapping inter-onset interval (s)* | 38 | 0.39 (0.19) |
| *Frequency over trial (Hz)* | 38 | 3.08 (1.07) |
| *Pointing target frequency over trial (Hz)* | 38 | 2.62 (0.66) |

**Abbreviations:** Hz: Hertz; N: Newton; s: Seconds

**Supplementary Table 3.** Rotated Component Loadings on the Q-motor Outcome Measures.

| **Task**<br>*Outcome measure* | **Q-motor Factor** |
| --- | --- |
| **Speeded tapping** |  |
| *Left finger; Area under the curve (N-s)* | **0.707** |
| *Right finger; Area under the curve (N-s)* | **0.723** |
| *Left finger; Inter-onset interval (s)* | **0.772** |
| *Right finger; Inter-onset interval (s)* | **0.859** |
| *Left foot; Inter-onset interval (s)* | **0.798** |
| *Right foot; Inter-onset interval (s)* | **0.893** |
| **Metronome tapping; Absolute deviation from pace (s)** |  |
| *Left finger; Paced; Fast* | **0.875** |
| *Left finger; Paced; Slow* | **0.832** |
| *Left finger; Unpaced; Fast* | **0.936** |
| *Left finger; Unpaced; Slow* | **0.913** |
| *Right finger; Paced; Fast* | **0.730** |
| *Right finger; Paced; Slow* | **0.672** |
| *Right finger; Unpaced; Fast* | **0.886** |
| *Right finger; Unpaced; Slow* | **0.865** |
| *Left foot; Paced; Fast* | **0.823** |
| *Left foot; Paced; Slow* | **0.883** |
| *Left foot; Unpaced; Fast* | **0.884** |
| *Left foot; Unpaced; Slow* | **0.900** |
| *Right foot; Paced; Fast* | **0.818** |
| *Right foot; Paced; Slow* | **0.885** |
| *Right foot; Unpaced; Fast* | **0.890** |
| *Right foot; Unpaced; Slow* | **0.740** |
| **Pointing (dominant hand)** |  |
| *Target frequency over trial (Hz)* | **-0.525** |
| **Pointing (dominant) and tapping (non-dominant)** |  |
| *Finger tapping area under the curve (N-s)* | **0.734** |
| *Finger tapping inter-onset interval (s)* | **0.788** |
| *Frequency over trial (Hz)* | **-0.756** |
| *Pointing target frequency over trial (Hz)* | **-0.568** |

All loadings were significant (>0.5 and < -0.5) and are highlighted in bold.
**Abbreviations:** Hz: Hertz; N: Newton; s: Seconds

**Supplementary Table 4.** Descriptive statistics and non-parametric pairwise comparisons for SANDI indices (significant for total HD sample vs HC) between HD-ISS 0-1, HD-ISS 2-3, and Healthy Controls.

| Microstructural variable | Region of interest | Descriptive statistics<br>Mean (SD) |  |  | Pairwise comparisons<br>Statistic, (p-value)<br>Effect size [CI 95%] |  |  |
| --- | --- | --- | --- | --- | --- | --- | --- |
|  |  | HD-ISS 0-1<br>N=13 | HD-ISS 2-3<br>N=17 | HC<br>N=57 | HD-ISS 0-1 vs HC | HD-ISS 2-3 vs HD-ISS 0-1 | HD-ISS 2-3 vs HC |
| Apparent soma density | Caudate | 0.45 (0.02) | 0.41 (0.07) | 0.47 (0.02) | 244 (0.057)<br>-0.228 [-0.43 – -0.02] | **56 (0.024)**<br>**-0.416 [-0.70 – -0.10]** | **119 (<0.001)**<br>**-0.546 [-0.69 – -0.38]** |
|  | Putamen | 0.42 (0.03) | 0.38 (0.05) | 0.43 (0.03) | 294 (0.251)<br>-0.138 [-0.35 – -0.01] | **53 (0.017)**<br>**-0.439 [-0.70 – -0.10]** | **174 (<0.001)**<br>**-0.464 [-0.64 – -0.25]** |
|  | Pallidum | 0.22 (0.03) | 0.19 (0.03) | 0.24 (0.04) | **229 (0.033)**<br>**-0.255 [-0.45 – -0.04]** | 64 (0.054)<br>-0.355 [-0.69 – -0.03] | **158 (<0.001)**<br>**-0.488 [-0.65 – -0.30]** |
| Apparent soma size | Caudate | 9.72 (0.17) | 9.88 (0.28) | 9.55 (0.13) | **587 (0.001)**<br>**0.391 [0.14 – 0.57]** | 150 (0.103)<br>0.302 [0.02 – 0.62] | **842 (<0.001)**<br>**0.534 [0.32 – 0.69]** |
|  | Putamen | 9.62 (0.19) | 9.77 (0.30) | 9.41 (0.14) | **597 (<0.001)**<br>**0.409 [0.19 – 0.60]** | 146 (0.143)<br>0.271 [0.02 – 0.60] | **832 (<0.001)**<br>**0.519 [0.27 – 0.68]** |
|  | Pallidum | 8.79 (0.52) | 8.11 (1.44) | 8.96 (0.51) | 263 (0.106)<br>-0.194 [-0.39 – -0.01] | **59 (0.033)**<br>**-0.394 [-0.69 – -0.06]** | **157 (<0.001)**<br>**-0.489 [-0.66 – -0.31]** |
| Extracellular signal fraction | Caudate | 0.37 (0.01) | 0.38 (0.03) | 0.36 (0.01) | **638 (<0.001)**<br>**0.483 [0.31 – 0.62]** | 148 (0.121)<br>0.287 [0.02 – 0.64] | **769 (<0.001)**<br>**0.425 [0.16 – 0.63]** |
|  | Putamen | 0.35 (0.01) | 0.37 (0.04) | 0.33 (0.01) | **597 (<0.001)**<br>**0.409 [0.22 – 0.56]** | 147 (0.132)<br>0.279 [0.02 – 0.62] | **776 (<0.001)**<br>**0.435 [0.17 – 0.64]** |
|  | Pallidum | 0.35 (0.04) | 0.38 (0.04) | 0.33 (0.02) | **517 (0.027)**<br>**0.264 [0.03 – 0.49]** | 157 (0.054)<br>0.355 [0.04 – 0.67] | **881 (<0.001)**<br>**0.592 [0.43 – 0.71]** |
| Extracellular diffusivity | Caudate | 1.39 (0.09) | 1.58 (0.24) | 1.42 (0.09) | 298 (0.277)<br>-0.131 [-0.36 – -0.01] | **169 (0.015)**<br>**0.447 [0.11 – 0.72]** | **697 (0.006)**<br>**0.317 [0.06 – 0.55]** |
|  | Putamen | 1.47 (0.12) | 1.63 (0.17) | 1.42 (0.09) | 490 (0.072)<br>0.216 [0.02 – 0.44] | **169 (0.015)**<br>**0.447 [0.13 – 0.71]** | **842 (<0.001)**<br>**0.534 [0.36 – 0.67]** |
|  | Pallidum | 1.67 (0.19) | 1.80 (0.18) | 1.53 (0.11) | **569 (0.003)**<br>**0.358 [0.09 – 0.56]** | 156 (0.060)<br>0.348 [0.04 – 0.65] | **893 (<0.001)**<br>**0.610 [0.47 – 0.72]** |

Abbreviations: CI, Confidence Intervals.
Pairwise comparisons were conducted with Mann–Whitney U tests; effect sizes are rank–biserial correlations (signed) with 95% CIs.
Bold indicates $p < 0.05$ .

